# Global spatiotemporal trends and determinants of COVID-19 vaccine acceptance on Twitter: a multilingual deep learning study in 135 countries and territories

**DOI:** 10.1101/2022.11.14.22282300

**Authors:** Xinyu Zhou, Xu Zhang, Heidi J. Larson, Alexandre de Figueiredo, Mark Jit, Samah Fodeh, Sten H. Vermund, Sujie Zang, Leesa Lin, Zhiyuan Hou

## Abstract

**Background:** COVID-19 vaccination has faced a range of challenges from supply-side barriers such as insufficient vaccine supply and negative information environment and demand-side barriers centring on public acceptance and confidence in vaccines. This study assessed global spatiotemporal trends in demand- and supply-side barriers to vaccine uptake using COVID-19-related social media data and explored the country-level determinants of vaccine acceptance.

**Methods:** We accessed a total of 13,093,406 tweets sent between November 2020 and March 2022 about the COVID-19 vaccine in 90 languages from 135 countries using Meltwater™® (a social listening platform). Based on 8,125 manually-annotated tweets, we fine-tuned multilingual deep learning models to automatically annotate all 13,093,406 tweets. We present spatial and temporal trends in four key spheres: (1) COVID-19 vaccine acceptance; (2) confidence in COVID-19 vaccines; (3) the online information environment regarding the COVID-19 vaccine; and (4) perceived supply-side barriers to COVID-19 vaccination. Using univariate and multilevel regressions, we evaluated the association between COVID-19 vaccine acceptance on Twitter® and (1) country-level characteristics regarding governance, pandemic preparedness, trust, culture, social development, and population demographics; (2) country-level COVID-19 vaccine coverage; and (3) Google® search trends on adverse vaccine events.

**Findings:** COVID-19 vaccine acceptance was high among Twitter® users in Southeast Asian, Eastern Mediterranean, and Western Pacific countries, including India, Indonesia, and Pakistan. In contrast, acceptance was relatively low in high-income nations like South Korea, Japan, and the Netherlands. Spatial variations were correlated with country-level governance, pandemic preparedness, public trust, culture, social development, and demographic determinants. At the country level, vaccine acceptance sentiments expressed on Twitter® predicted higher vaccine coverage. We noted the declining trend of COVID-19 vaccine acceptance among global Twitter® users since March 2021, which was associated with increased searches for adverse vaccine events. **Interpretation** In future pandemics, new vaccines may face the potential low-level and declining trend in acceptance, like COVID-19 vaccines, and early responses are needed. Social media mining represents a promising surveillance approach to monitor vaccine acceptance and can be validated against real-world vaccine uptake data.

**Funding:** National Natural Science Foundation of China.

## Introduction

Widespread distribution, acceptance, and uptake of SARS-CoV-2 (COVID-19) vaccines are crucial to reducing the severity and transmissibility of SARS-CoV-2 infections. During the first year (2021) of global vaccination efforts, COVID-19 vaccines saved an estimated 19.8 million lives.^1^ However, COVID-19 vaccination faced both supply-side barriers such as insufficient vaccine supply and negative information environment and demand-side barriers centring on public acceptance and confidence in vaccines. In past decades, vaccine hesitancy – the delay in acceptance or refusal of vaccines despite their availability^2^ – had been prevalent across the world. A better understanding of COVID-19 vaccine acceptance and hesitancy is crucial for pandemic control.

Most previous studies on COVID-19 vaccine acceptance are surveys.^3-5^ Surveys have found high heterogeneity in vaccine acceptance across nations, ranging from more than 90% (e.g., Vietnam, India, and China) to around 40% (e.g., France, Serbia, and Croatia)^3,4^. Serial cross-sectional surveys help track the state of vaccine acceptance among the public, but costs limit generalizable longitudinal global assessments.

A rapidly growing field is the use of social media “big data” and machine learning to conduct behavioural surveillance, using social media content to understand public attitudes towards vaccines and other health knowledge, attitudes, and behaviours.^6,7^ Although social media users are a subgroup of the public, spatiotemporal trends of their attitudes could examine how the global and local social media atmospheres reshape mindsets over time. Previous studies have used data from social media platforms such as Twitter® and Facebook® to analyse COVID-19 vaccine acceptance, principally from high-income countries such as the US, UK, and Canada.^8-13^ Trends in vaccine attitudes in low- and middle-income countries (LMICs) are less well elucidated.

We performed social media surveillance on COVID-19 vaccine acceptance using machine learning techniques. By analyzing millions of multilingual tweets on COVID-19 vaccination from late 2020 to early 2022, we sought to assess the global spatiotemporal trends in the perceived demand- and supply-side barriers to COVID-19 vaccine uptake among Twitter® users in 135 countries and territories, covering vaccine acceptance and confidence, online information environment, and perceived barriers to accessing vaccines. We further investigated the determinants of vaccine acceptance as expressed on Twitter® and assessed the associations between vaccine acceptance on Twitter® and vaccine coverage. Our study provides a new and robust surveillance approach to timely and early monitor health risks, public attitudes, and behaviours during pandemics. This can be leveraged as a supplement to existing public health surveillance approaches in addressing global health issues.

## Methods

The method section is written for readers without a machine learning background, and methodological details were presented in Supplementary Material 1. All data analyses/visualization were performed using <u>Python</u> (version 3.7.2) and <u>R</u> (version 4.2.1).

### Collection of COVID-19 vaccine-related tweets

As one of the world’s most popular social media platforms, Twitter® has 217 million daily active users globally and was used as our data source.^14^ We hired professional translators from a translation company (Beijing Chinese-Foreign Translation & Information Service Co., Ltd.) and leveraged <u>Google Translate</u> to verify 1,027 multilingual keywords on the COVID-19 vaccine in 90 major languages on Twitter®. We present a flowchart of keyword identification in the supplementary material (Figure S1) and our final keywords in the supplementary worksheet Table #1. Using the multilingual keywords identified, we collected all publicly-available tweets (n=13,093,406) from Twitter® on COVID-19 vaccination from November 13, 2020, to March 5, 2022, through the <u>Meltwater®</u> media monitoring and social listening platform.

### Manual annotation of sampled tweets

To leverage deep learning models for auto-annotating tweets, a common practice is to annotate tweets first by humans and then train the deep learning model to imitate human annotation. According to the framework of vaccine hesitancy proposed by the WHO,^2^ we developed an annotation framework for COVID-19 vaccine-related tweets and validated it using a 500-tweet subsample. In this framework, vaccine acceptance or hesitancy is the core measure, which is determined by vaccine confidence, information environment, and vaccination convenience (supply-side barriers). Based on the annotation framework, we manually annotated 8,125 English-language tweets on COVID-19 vaccination. Each tweet was annotated by two annotators independently, and a third annotator resolved any disagreements.

The annotation framework has two major steps: First, annotators filtered human-generated tweets from those generated by bot accounts, news reports, advertisements, and governmental announcements; which were removed and archived. Second, the human-generated COVID-19 vaccine-related tweets were annotated according to their relevance to the eight predefined categories in our annotation framework. The framework includes eight categories in four key spheres: (1) COVID-19 vaccine acceptance, covering two categories - intent to accept vaccination and intent to reject vaccination; (2) confidence in COVID-19 vaccines, covering three categories - belief that the vaccines are effective, belief that the vaccines are not safe, and distrust in government; (3) the online information environment regarding the COVID-19 vaccine - misinformation or rumours about the vaccines; and (4) perceived supply-side barriers to COVID-19 vaccination, covering two categories - vaccine accessibility and vaccine distribution. The definition of each COVID-19 vaccine category is available in Table S2. Depending on their contents, tweets could be annotated into one category, multiple categories, or no category.

### Deep learning annotation of all multilingual tweets

To extend our manual classifications (n=8,125), we used XLM-RoBERTa (XLM-R), a deep learning model with state-of-the-art performance in cross-lingual understanding benchmarks.^15^ XLM-R is a large multi-lingual language model, pretrained on 100 different languages using more than two terabytes of filtered CommonCrawl data; this model leads to significant performance gains for a wide range of cross-lingual transfer tasks. ^15^ To analyze new multilingual texts, it only needs to be finetuned with a small-sample mono-lingual dataset.^15^ Therefore, we finetuned the XLM-R model using the aforementioned manually-annotated English-language tweets dataset to automatically analyze the balance of the 13,093,406 COVID-19 vaccine-related tweets.

To finetune the XLM-R model, we randomly selected 80% of our annotated tweets as the training set, 10% as the validation set, and 10% as the test set. The training set allowed the models to learn how to annotate tweets to follow the guidance of the prior human annotators. The validation set enabled us to find proper hyperparameters for the models to learn. In the test set, we evaluated the model’s performance. The deep learning models achieved accuracies from 72.7% to 89.7% in automatically identifying human-generated tweets and annotating them into the eight predefined categories in our annotation framework. The models were trained, validated, and tested on <u>AutoDL®</u>.

Finally, we used our fine-tuned XLM-R model to annotate all COVID-19 vaccine-related tweets (n=13,093,406) automatically. Like our manual annotation process, the model first identified if each tweet was sent by humans or not. The tweets which were deemed not to be sent by a human (n=6,046,183) were excluded by the model. The remaining human-generated tweets (n=7,047,223) were annotated by the deep learning model into the aforementioned eight categories. We present an overview of the data collection and analysis process in Figure 1.

**Figure 1.**
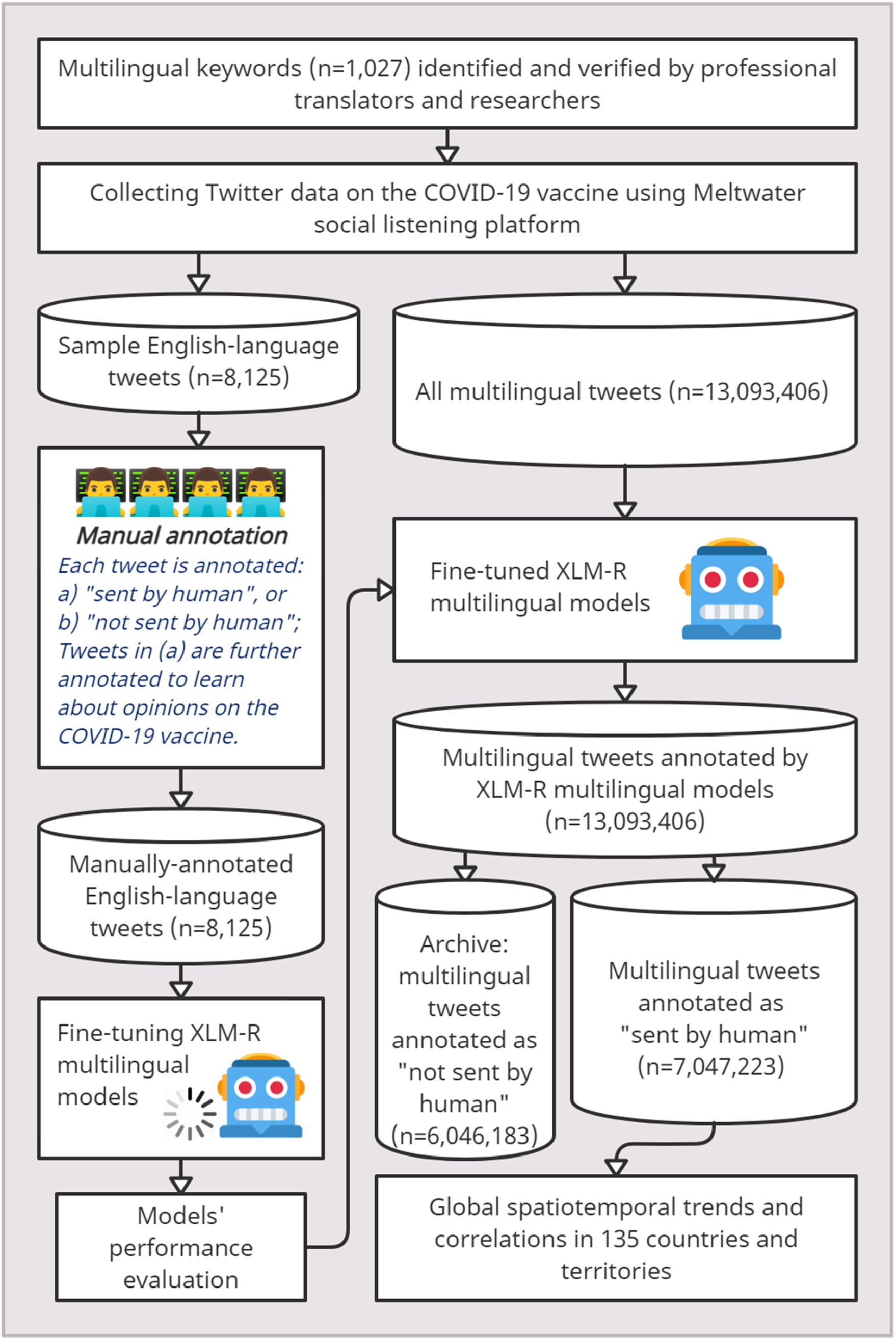
An overview of the methodology.

### Triangulating data and statistical analysis

Statistical analyses were based on the annotated results of the fine-tuned XLM-R model in determining the categories of all human-generated vaccine-related tweets. For each of the eight predefined categories in our annotation framework, tweets were aggregated by time and geo-location according to the model’s annotated results. We calculated the opinion of each Twitter user by averaging all his/her tweets within each time interval and then calculated the average opinions of Twitter® users spatiotemporally. All calculations were based on the subset of human-generated and geo-located tweets, except that global temporal trends were calculated on the subset of human-generated tweets.

First, spatial trends of COVID-19 vaccine acceptance and its determinants on Twitter® were estimated at the country level using only tweets with geolocation. We focused on countries and territories meeting the following criteria: (1) Twitter® is not banned by the relevant government, (2) there are data from at least 100 Twitter® users in that country or territory, (3) the analyzed 90 languages cover all official languages in that country or territory. There were 171 countries and territories with tweets sent by at least 100 Twitter® users, and their censorship status and official languages are shown in Supplement table #2. After excluding those with official languages not covered by our dataset and those banning Twitter®, we included 135 countries or territories.

To explore the determinants of the spatial trends in COVID-19 vaccine acceptance on Twitter®, we first estimated their correlation with vaccine confidence, information environment, and perceived vaccine convenience on Twitter®. We further collected 20 country-level indicators regarding governance, pandemic preparedness, trust, culture, social development status, and demographics from multiple external sources (supplementary material Table S4) and evaluated their correlations with COVID-19 vaccine acceptance on Twitter® using univariant linear regression.

To explore the public health consequences of COVID-19 vaccine acceptance on Twitter®, we linked country-level acceptance to coverage of COVID-19 vaccination using univariant linear regression. We further explored the linkages using multilevel regression to take into account the geographic regions and country income levels.

Second, global temporal trends are calculated at the daily level. Spline regression was employed to fit global temporal trends in opinions of the COVID-19 vaccine on Twitter®. Country-level trends were calculated at the weekly and monthly levels in countries with sufficient data.

To explain the temporal trends in COVID-19 vaccine acceptance on Twitter®, we obtained weekly country-level Google® Search Trends data on 12 topics relating to adverse events following immunisation (AEFI). The 12 topics were selected based on our team’s knowledge about AEFI and verified using the five most relevant topics and queries according to Google® Trends (Supplement Worksheet #4). We utilized univariant linear regression to explore their associations to weekly-level trends of COVID-19 vaccine acceptance on Twitter® in each country.

### Role of funding

The funders have no role in study design, data analysis and interpretation of data, the writing of the manuscript, or the decision to submit the paper for publication.

## Results

Globally, we identified and annotated 7,047,223 multilingual human-generated tweets on the COVID-19 vaccine between November 13, 2020, and March 5, 2022 (sent by 3,344,144 Twitter® users); of which 58.7% (4,137,550) had standardised geo-location data (sent by 1,953,157 users). On average, each Twitter® user sent fewer than two tweets relating to the COVID-19 vaccine (Table S5). Among geo-located tweets, the United States accounted for the largest proportion (1,792,206, 43.3%), followed by the United Kingdom (368,037, 8.9%), Canada (248,252, 6.0%), Japan (196,502, 4.8%), and India (164,506, 4.0%). During the study period, the number of tweets increased significantly in December 2020, as the first COVID-19 vaccine – the Pfizer/BioNTech® mRNA vaccine was authorized and released, followed by a downward trend beginning in January 2021 (Supplementary Material Figure S2a).

### Spatial variation of COVID-19 vaccination-related opinions expressed on Twitter

Figures 2 and S3 present the spatial variation of COVID-19 vaccination-related opinions of Twitter® users from 135 countries or territories, including 12 from the Western Pacific Region (WPR), six from the Southeast Asian Region (SEAR), 41 from the European Region (EUR), 18 from Eastern Mediterranean Region (EMR), 31 from Americas Region (AMR), and 27 from African Region (AFR). The COVID-19 vaccine acceptance varied across the WHO regions. Tweets from SEAR, EMR and WPR demonstrated higher COVID-19 vaccine acceptance and expressed more confidence in vaccine effectiveness and safety than tweets from AMR, EUR and AFR. SEAR dominated the top 10 countries or territories with high vaccine acceptance (four out of 10 countries, Bangladesh, 78.1%; India, 68.0%; Nepal, 66.0%; Indonesia, 64.3%). Of the 10 countries or territories with the highest proportion of Twitter® users intending to refuse COVID-19 vaccines, four were from AMR (Guadeloupe, 24.4%; Martinique, 21.1%; Haiti, 20.2%; Argentina, 19.6%), and four were from AFR (Réunion, 24.1%; Gabon, 22.2%; the Democratic Republic of the Congo, 20.8%; Republic of the Congo, 19.7%).

**Figure 2.**
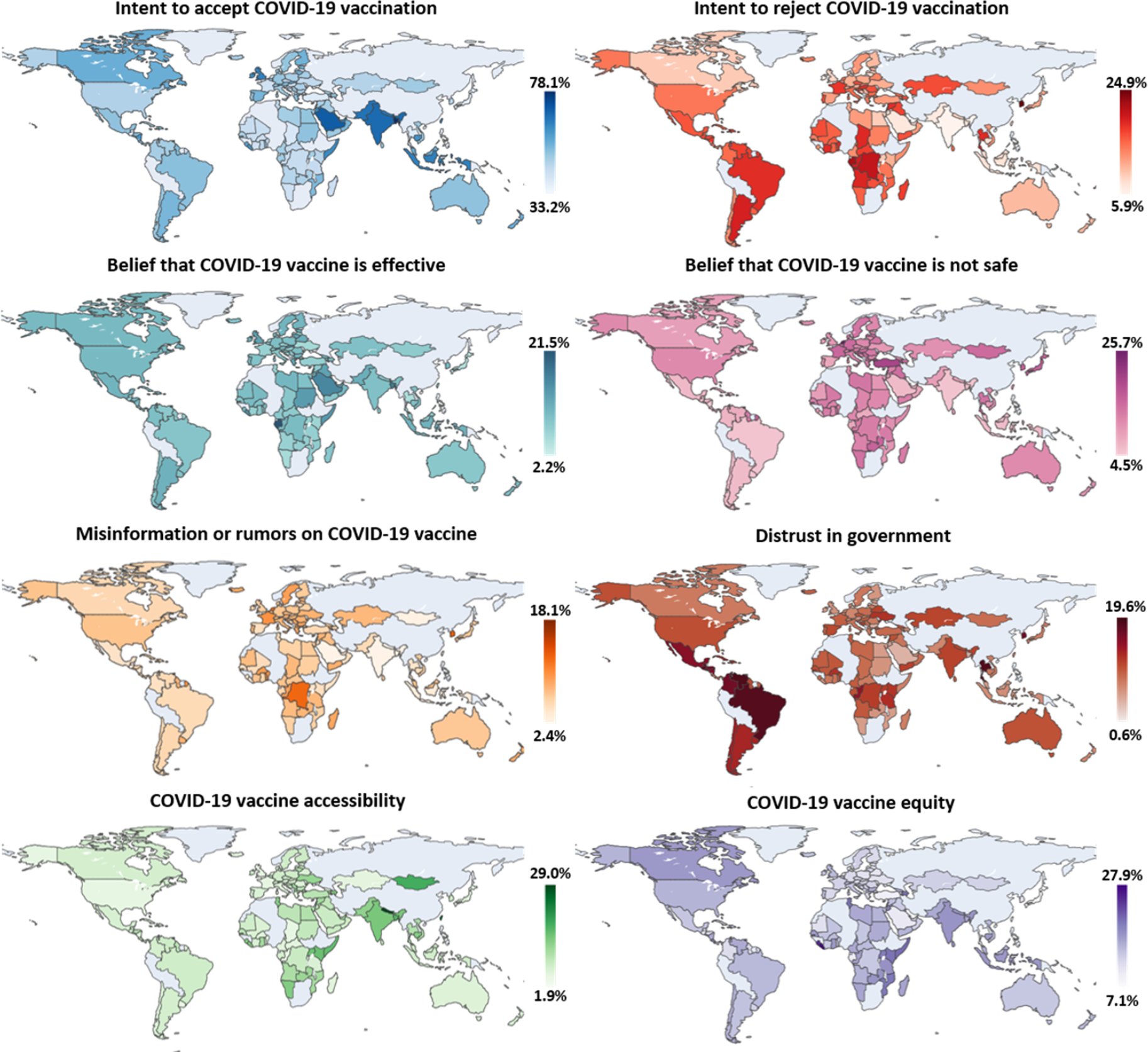
Geographical variations of COVID-19 vaccine acceptance, vaccine confidence, information environment, and prevalence of vaccine access-related discourse among Twitter users, November 13, 2020 – March 5, 2022 (n=135 countries or territories). • The graphs below illustrate the proportion of Twitter users expressing each opinion regarding COVID-19 vaccination. • Only countries or territories with COVID-19 vaccination-related tweets from >100 Twitter users are presented. • Countries or territories with inadequate data or government blocking the access to Twitter, and those not covered by 90 common languages are in grey.

The proportion of tweets indicating a belief that the COVID-19 vaccine is unsafe was higher in the Netherlands (25.7%), Reunion (20.7%), and Turkey (19.7%) than in other countries/territories. Countries from AMR had the lowest proportion of tweets indicating a belief that the COVID-19 vaccine is unsafe (six out of 10 countries: Nicaragua, 4.5%; Grenada, 4.9%; Brazil, 5.2%; Uruguay, 5.4%; EI Salvador, 5.8%; Guatemala, 5.9%), while professing more distrust in the government (eight of the top 10 countries: El Salvador, 19.6%; Venezuela, 18.4%; Brazil, 18.0%, Guatemala, 17.8%; Colombia, 16.5%; Honduras, 16.2%; Nicaragua, 16.0%; Mexico, 15.2%) and presenting more misinformation/rumours (four of the top 10 countries: Guadeloupe, 18.1%; Martinique, 15.5%; Saint Kitts And Nevis, 14.3%; Cayman Islands, 11.3%). Twitter® users in SEAR (Nepal, 29.0%; Bangladesh, 17.7%; India, 15.0%) and AFR (Kenya, 16.1%; Uganda, 15.9%; Liberia, 14.7%) discussed COVID-19 vaccine accessibility most frequently. The proportion of tweets concerning COVID-19 vaccine equity was higher in AFR (five of the top 10 countries: Liberia, 27.9%; Sierra Leone, 25.2%; Kenya, 20.7%; Mozambique, 20.3%; Uganda, 20.0%) than in other regions.

### Country-level analysis of COVID-19 vaccine acceptance

Table 1 describes the country-level characteristics from Twitter® and external sources associated with COVID-19 vaccine acceptance among Twitter® users from univariate linear regressions. All six internal country-level indicators from Twitter® regarding vaccine confidence, online information environment, and perceived supply-side barriers to COVID-19 vaccination were strong predictors of vaccine acceptance and refusal among Twitter® users, indicating internal consistency. Among all external country-level characteristics analysed, only trust in government (R^2^=9.6%) and internet coverage (R^2^=10.6%) were significantly correlated with vaccine acceptance. On the other hand, almost all country-level characteristics were significantly correlated with vaccine refusal. Especially, better governance (government effectiveness, R^2^=6.8%; control of corruption, R^2^=6.8%) and preparedness for pandemics (epidemic ready score, R^2^=15.9%; global health security index, R^2^=4.7%) were negatively correlated with vaccine refusal, whereas state fragility (R^2^=9.3%) was positively correlated. Trust in government (R^2^=6.3%) was negatively linked to vaccine refusal, and a culture of uncertainty avoidance (R^2^=25.8%) was positively linked. Negative links to vaccine refusal were established for better social development status (socio-demographic index, R^2^=6.1%; GDP per capita, R^2^=3.3%; school enrollment, R^2^=3.3%; internet coverage, R^2^=7.8%). In terms of demographic characteristics, population density (R^2^=7.3%) was negatively linked to vaccine refusal, whereas the proportion of the population aged 0-14 (R^2^=4.9%) was positively linked.

**Table 1.** Associations between country-level characteristics and COVID-19 vaccine acceptance by univariate linear regressions (n=135 countries or territories). • Only countries or territories with COVID-19 vaccination-related tweets from >100 Twitter users were analysed.

| Variables | Intent to accept COVID-19 vaccination |  |  |  | Intent to reject COVID-19 vaccination |  |  |  |
| --- | --- | --- | --- | --- | --- | --- | --- | --- |
|  | Coefficient | 95% CI | P-value | R<sup>2</sup> | Coefficient | 95% CI | P-value | R<sup>2</sup> |
| *Vaccine-related discourse on Twitter* |  |  |  |  |  |  |  |  |
| Belief that COVID-19 vaccine is effective | 1.880 | [1.523,2.237] | <0.001 | 44.9% | -0.497 | [-0.733,-0.260] | <0.001 | 11.5% |
| Belief that COVID-19 vaccine is not safe | -1.351 | [-1.678,-1.440] | <0.001 | 33.5% | 0.376 | [0.176,0.575] | <0.001 | 9.4% |
| Misinformation or rumours on COVID-19 vaccine | -1.886 | [-2.332,-1.353] | <0.001 | 34.5% | 0.978 | [0.744,1.212] | <0.001 | 33.9% |
| Distrust in government | -0.591 | [-0.995,-0.187] | 0.004 | 5.9% | 0.547 | [0.351,0.744] | <0.001 | 18.6% |
| COVID-19 vaccine accessibility | 0.810 | [0.489,1.132] | <0.001 | 15.7% | -0.447 | [-0.613,-0.280] | <0.001 | 17.5% |
| COVID-19 vaccine equity | 0.667 | [0.255,1.079] | 0.002 | 7.2% | -0.243 | [-0.462,-0.023] | 0.031 | 3.5% |
| *Governance* |  |  |  |  |  |  |  |  |
| Government effectiveness | 0.182 | [-1.115,1.479] | 0.782 | 0.1% | -1.008 | [-1.660,-0.358] | 0.003 | 6.8% |
| Control of corruption | 0.098 | [-1.207,1.404] | 0.882 | 0.0% | -1.012 | [-1.667,-0.356] | 0.003 | 6.8% |
| State fragility | -0.124 | [-0.638,0.391] | 0.635 | 0.2% | 0.456 | [0.199,0.712] | 0.001 | 9.3% |
| *Pandemic preparedness* |  |  |  |  |  |  |  |  |
| Epidemic Ready Score | 0.061 | [-0.037,0.158] | 0.217 | 2.5% | -0.081 | [-0.129,-0.033] | 0.001 | 15.9% |
| Global Health Security Index | -0.006 | [-0.099,0.087] | 0.906 | 0.0% | -0.058 | [-0.105,-0.010] | 0.018 | 4.7% |
| Doctors per 1000 people | 0.001 | [-0.072,0.073] | 0.983 | 0.0% | -0.022 | [-0.06,0.016] | 0.257 | 1.1% |
| *Trust* |  |  |  |  |  |  |  |  |
| Trust in government, % | 0.127 | [0.039,0.216] | 0.005 | 9.6% | -0.056 | [-0.105,-0.007] | 0.025 | 6.3% |
| Trust in science, % | 0.057 | [-0.081,0.196] | 0.413 | 0.8% | -0.024 | [-0.096,0.048] | 0.504 | 0.6% |
| Interpersonal trust, % | -0.059 | [-0.246,0.129] | 0.530 | 1.0% | -0.036 | [-0.135,0.062] | 0.460 | 1.4% |
| *Culture-related index* |  |  |  |  |  |  |  |  |
| Individualism | 0.002 | [-0.086,0.089] | 0.973 | 0.0% | -0.027 | [-0.073,0.018] | 0.235 | 2.6% |
| Uncertainty avoidance | -0.068 | [-0.156,0.021] | 0.131 | 4.3% | 0.088 | [0.047,0.129] | <0.001 | 25.8% |
| *Social development status* |  |  |  |  |  |  |  |  |
| Socio-Demographic Index | 0.419 | [-6.830,7.668] | 0.909 | 0.0% | -5.268 | [-8.911,-1.625] | 0.005 | 6.1% |
| Ln (GDP per capita) | -0.134 | [-1.048,0.780] | 0.772 | 0.1% | -0.474 | [-0.941,-0.007] | 0.047 | 3.3% |
| School enrollment, % | 0.005 | [-0.035,0.045] | 0.806 | 0.0% | -0.021 | [-0.042,0.001] | 0.042 | 3.3% |
| Internet coverage, % | 0.140 | [0.024,0.257] | 0.020 | 10.6% | -0.067 | [-0.133,-0.001] | 0.048 | 7.8% |
| *Demographics* |  |  |  |  |  |  |  |  |
| Population density | 0.001 | [-0.000,0.003] | 0.144 | 1.7% | -0.001 | [-0.002,-0.001] | 0.002 | 7.3% |
| Population ages 0-14, % | -0.029 | [-0.157,0.099] | 0.654 | 0.2% | 0.085 | [0.018,0.151] | 0.013 | 4.9% |
| Population ages 65 and above, % | -0.160 | [-0.343,0.023] | 0.087 | 2.4% | -0.048 | [-0.146,0.050] | 0.338 | 0.7% |
| Urban population, % | 0.008 | [-0.054,0.071] | 0.790 | 0.1% | -0.011 | [-0.044,0.022] | 0.508 | 0.3% |

We further evaluated the associations between Twitter® users’ acceptance and real coverage of COVID-19 vaccination across the country using univariate linear regressions and multilevel regressions (Figure 3, Figure S4, and Table S6). We found that vaccination acceptance expressed on Twitter® was a predictor of COVID-19 vaccine coverage at the country level, and we noted similar associations across time, place, and income groups.

**Figure 3.**
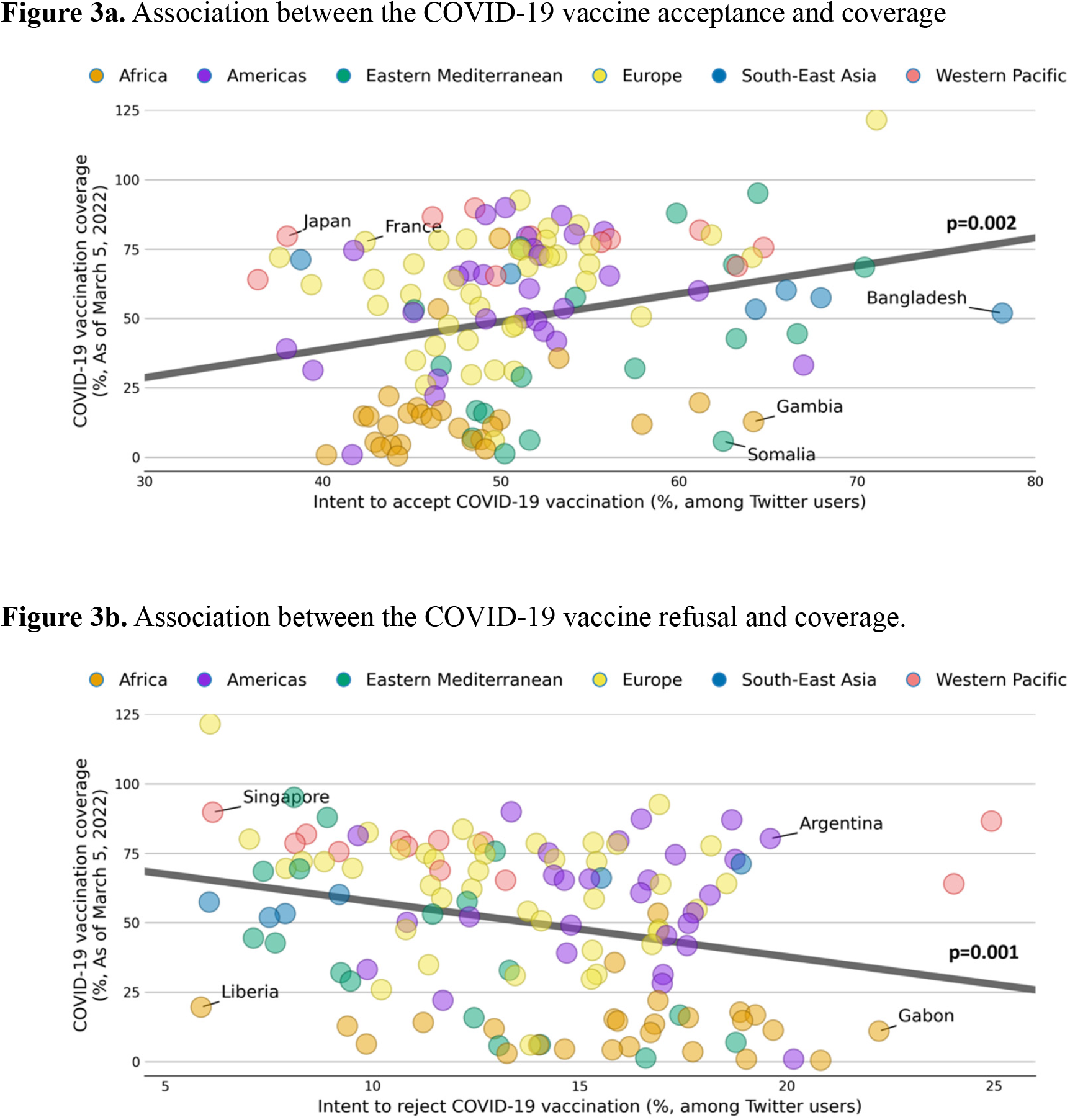
Association between Twitter users’ acceptance and real coverage of COVID-19 vaccination across 131 countries and territories. • Four countries had no data on COVID-19 vaccine coverage and were excluded.

### Temporal trends of COVID-19 vaccination-related opinions expressed on Twitter

Figure 4a shows the temporal trends of COVID-19 vaccination-related opinions on Twitter® globally. Among Twitter® users commenting on COVID-19 vaccines, the proportion of those indicating their acceptance of COVID-19 vaccination increased from a daily average of 44.1% in December 2021 to 56.0% in March 2021 and then began a slow decline, reaching a daily average of 33.4% between February 1 and March 5, 2022. The intent to refuse COVID-19 vaccination slowly climbed from a daily average of 10.5% in February 2021 to 17.2% in August 2021, and then kept stable. The proportion of Twitter users believing in the COVID-19 vaccine’s effectiveness first rose to around 11.6% in March 2021 and then slowly declined to around 5%. Meanwhile, the proportion of Twitter® users expressing their disbelief in the COVID-19 vaccine’s safety went through three significant upward phases from March 2021 (9.6%) to June 2021 (13.6%), August 2021 (12.4%) to October 2021 (16.6%), and December 2021 (17.5%) to February 2022 (18.9%). The proportion of people who expressed distrust in governments decreased throughout 2021, yet increased slightly during early 2022, from December 2021 (10.8%) to February 2022 (12.2%). Misinformation and rumours regarding COVID-19 vaccines generally increased during the observation period, with a notable increase between March 2021 (4.3%) and August 2021 (10.4%). The proportion of Twitter® users discussing COVID-19 vaccine accessibility remained largely stable (8.5%) until June 2021, when it began to decline gradually, reaching a daily average of 2.7% in January 2022. Meanwhile, discourse concerning COVID-19 vaccine equity gradually increased in the second half of 2021, reaching 17.4% in August 2021 and then keeping stable. Temporal trends at the regional level are similar to global trends (Figure S5).

**Figure 4.**
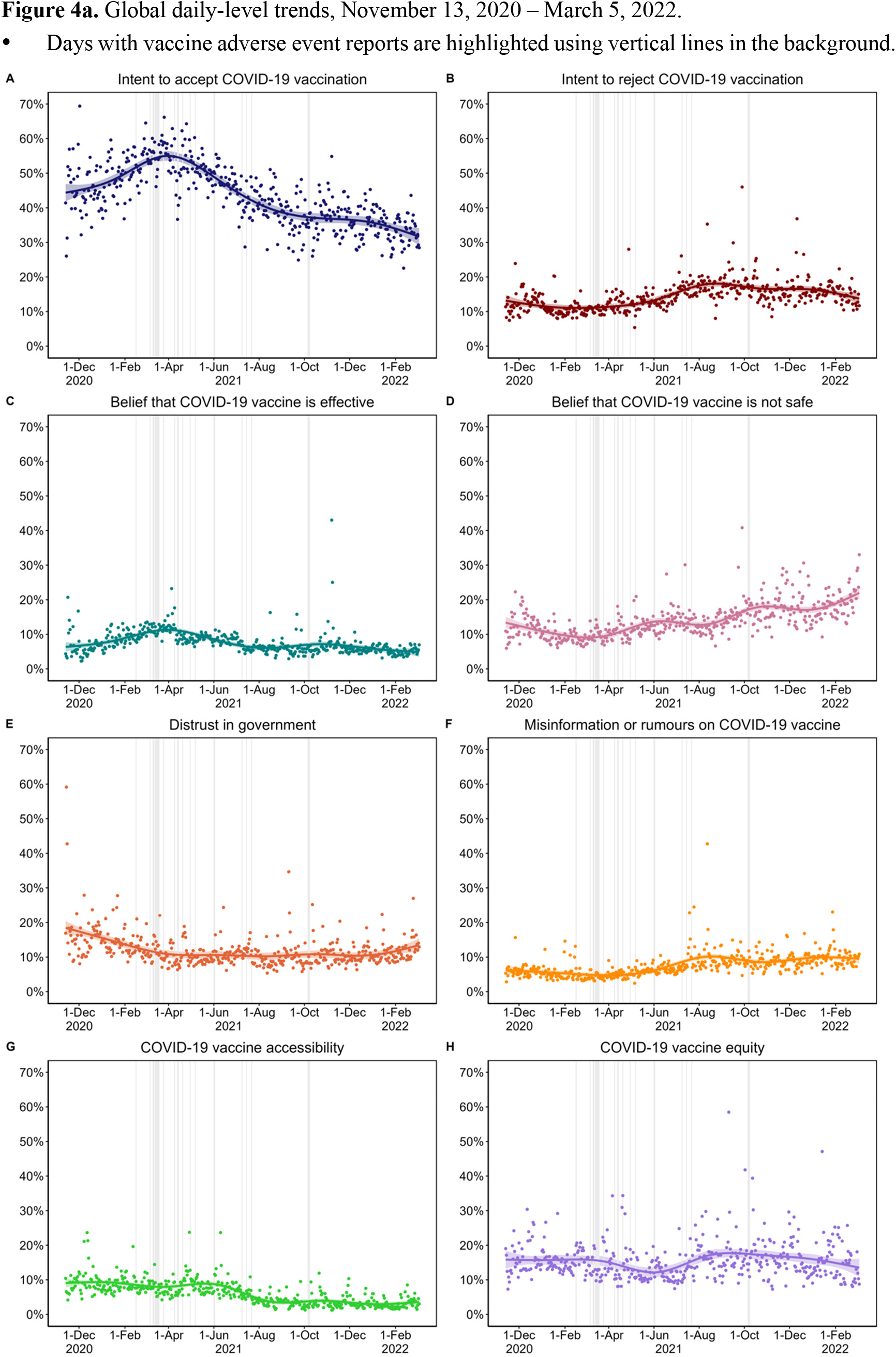
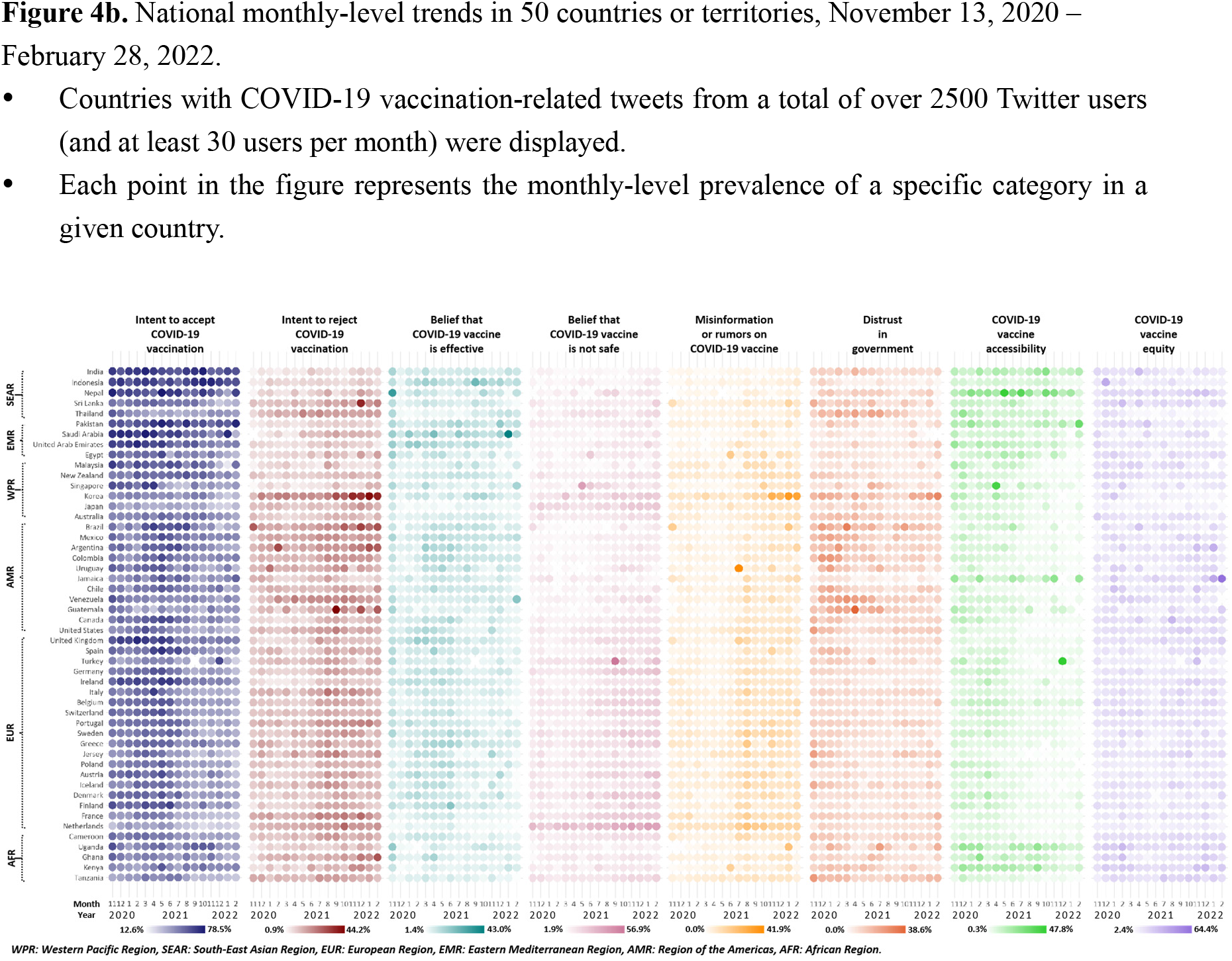
Global temporal trends of weighted COVID-19 vaccine acceptance, vaccine confidence, information environment, and prevalence of vaccine access-related discourse among Twitter users.

Noted that the time frame when COVID-19 vaccine acceptance on Twitter® began to decline coincided with the global reports on AEFI of the COVID-19 vaccine, and similar findings were found in vaccine refusal, vaccine confidence, and information environment.

Figure 4b shows the country-level temporal/monthly trends of COVID-19 vaccination-related opinions on Twitter® for 50 countries with sufficient numbers of tweets. During November 2020 and February 2022, most countries, especially countries in AMR and EUR, witnessed a slight initial rise in COVID-19 vaccine acceptance followed by a sustained decline, while the prevalence of COVID-19 vaccination refusal increased. In most countries, especially in AMR and EUR, a decreasing proportion of Twitter® users expressed their belief in the COVID-19 vaccine’s effectiveness, or distrust in governments, and discussed COVID-19 vaccine accessibility. Meanwhile, an increasing proportion of Twitter® users sent misinformation or rumours and expressed concerns about the COVID-19 vaccine safety. However, temporal trends were diverse and inconsistent in some countries. Among Twitter® users in SEAR and EMR countries, such as India, Indonesia, and Pakistan, COVID-19 vaccine acceptance and confidence remained high, whereas South Korea, Japan, and the Netherlands had persistently low acceptance and confidence in the COVID-19 vaccine.

### Association between temporal trends of vaccine acceptance and AEFI-related search on Google

The year of 2021 observed numerous reports on COVID-19 vaccine AEFI (supplementary material 2 worksheet #3), triggering an unprecedented volume of vaccine adverse event-related searches on Google® worldwide (Figure S7). We identified 12 AEFI-related topics on Google® and collected the weekly country-level search trends (Figure S8). We then estimated the weekly relationships between AEFI Google® Trends and COVID-19 vaccine acceptance or refusal on Twitter® in each of the 26 countries with sufficient tweets by univariate linear regressions. Totally we ran 620 regressions, of which 229 (36.9%) significant correlations were noted (Figure 5). These regression coefficients over time revealed that the later an AEFI-related search peaked, the more likely it was linked to a decline in COVID-19 vaccine acceptance and an increase in refusal on Twitter® (p<0.001). Consistent findings were identified for most AEFI-related topics.

**Figure 5.**
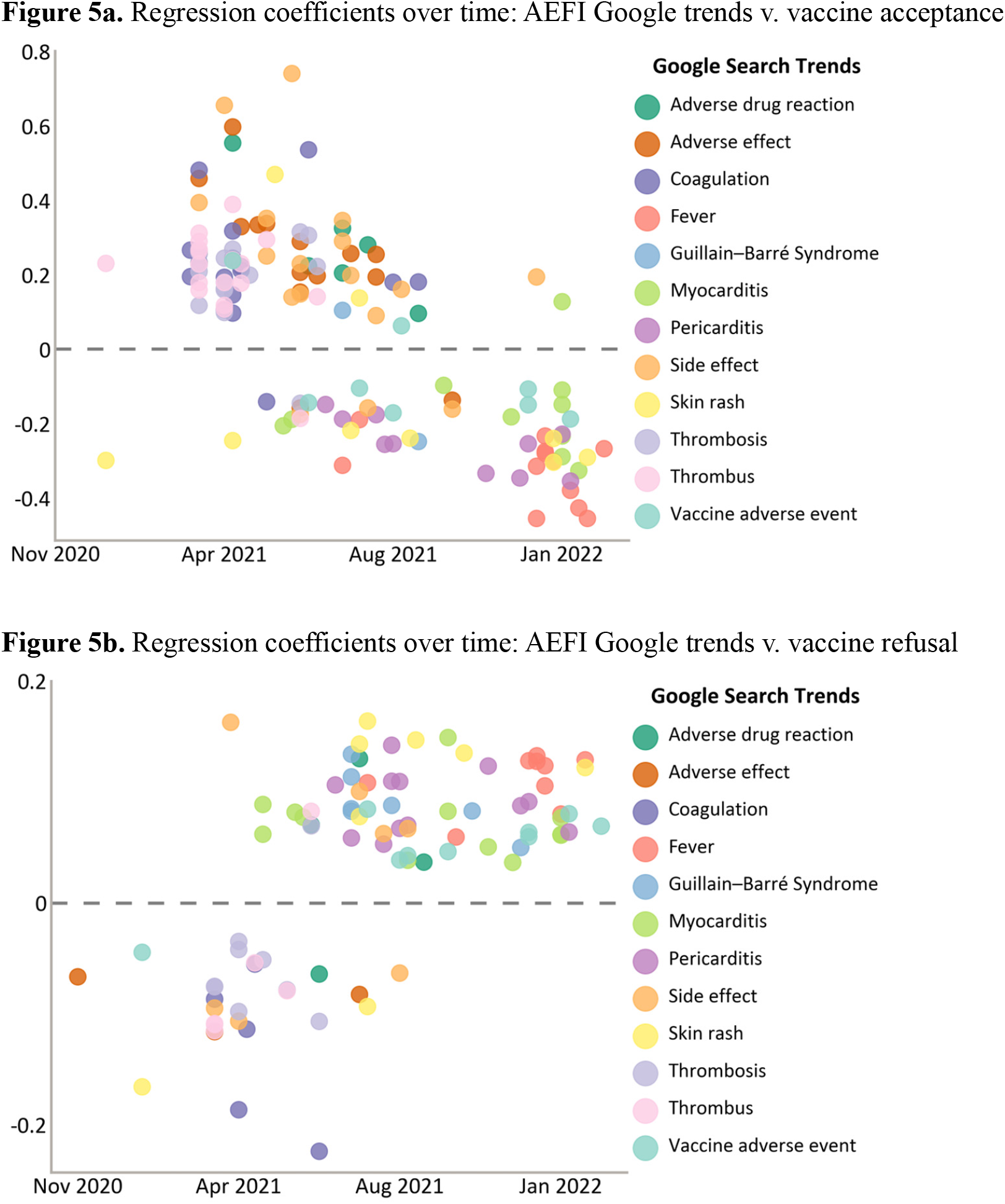
Temporal variation of the association between COVID-19 vaccine acceptance on Twitter and AEFI Google search Trends, November 13, 2020 – February 20, 2022. • Only the 26 countries or territories with both sufficient Twitter data (total number of Twitter users mentioning COVID-19 vaccination >5000; weekly number >28) and Google Trends data were analysed. No analysis after February 21, 2022, due to data insufficiency. • We linked weekly-level AEFI Google Trends data to weekly-level COVID-19 vaccine acceptance on Twitter in each of the 26 countries to calculate the regression coefficients. Totally 620 univariant linear regressions were run (4 with missing data were not run), of which 229 (36.9%) were significant (p<0.05) and their regression coefficients are shown. • The y-axis is the regression coefficient, and the x-axis is the time when the country-level Google search trends on AEFI peaked.

## Discussion

We monitored spatiotemporal trends of discourse regarding COVID-19 vaccination among global, multilingual Twitter® users in 135 countries or territories, covering the time from emergency use approval of COVID-19 vaccines in diverse settings to the time when over half of the world population had been vaccinated. Our estimation of COVID-19 vaccine acceptance from Twitter® can predict real-world vaccine coverage worldwide. We found that COVID-19 vaccine acceptance was higher among Twitter® users in Southeast Asia and Eastern Mediterranean Regions than in the rest of the world. Spatial variations were correlated with country-level governance, pandemic preparedness, public trust, culture, social development, and demographic determinants. In most countries, there was a declining trend in COVID-19 vaccine acceptance and confidence among Twitter® users as AEFI spread. AEFI-related searches online were increasingly linked to lower vaccine acceptance and higher refusal in diverse countries and territories.

We performed social media monitoring based on multilingual deep learning models, which can be leveraged as a supplement to existing public health surveillance approaches in addressing global health issues mediated by service availability or health-seeking behaviours.^16,17^ As a supplement to traditional approaches such as surveys, social media-based studies could identify regions or sub-populations with low COVID-19 vaccine acceptance close to real-time and emerging concerns to early warn vaccine-related risks or crisis, which help to inform timely and targeted interventions. It also offers opportunities to improve global equity in research capacity and pandemic response because it is highly favourable for settings with limited budgets and urgent timelines. Future studies should explore the vast potential of social media monitoring and how it can provide surveillance data to address a wide array of global health challenges.

We provide international evidence regarding the linkage between COVID-19 vaccine acceptance on Twitter and vaccine uptake behaviour. This finding highlights the potential public health consequences of social media interventions as well as the surveillance potential of social media monitoring. With the popularity of social media, there could be an “infodemic” during a pandemic, which refers to the overwhelming information and misinformation mainly on social media that could have population health consequences.^18^ Social media monitoring studies enable public health scientists, authorities, and policymakers to provide “infodemic” surveillance and enable social media-based or other interventions. Online interventions such as chatbots and videos containing prosocial and altruistic messages have proven to be viable interventions to promote vaccine acceptance. ^19,20^ Given the pervasive penetration of social media, these strategies will be increasingly important.

Through spatial analysis, we found that COVID-19 vaccine acceptance expressed on Twitter® varied from 33.2% to 78.1% while vaccine refusal varied from 5.9% to 24.9% across countries. The acceptance and confidence in the COVID-19 vaccine were relatively high in Southeast Asia but low in Africa and America. Our findings are consistent with previous global surveys on vaccine acceptance. ^21-24^ It is possible that biases in who responds to surveys and the non-randomness of social media users may not result in highly diverse findings. All social development indicators were found to be negatively associated with vaccine refusal. Africa faced dual challenges in insufficient supply and low acceptance of COVID-19 vaccines and should be paid more attention. ^3^

Tweets suggest that better governance, greater pandemic preparedness, and greater trust in government might reduce COVID-19 vaccine refusals and increase vaccine coverage. In countries with higher COVID-19 vaccine refusal rates, such as El Salvador, Venezuela, and Thailand, tweets indicated deeper distrust in the government. These results support previous research that suggested higher levels of trust and less government corruption to be significantly associated with higher COVID-19 vaccine coverage, leading to lower COVID-19 infection rates.^25^ A 19-country survey also revealed that countries with high COVID-19 vaccine acceptance tended to be those with strong trust in the central government.^4^ Trust is associated with compliance with public health guidance such as mask-wearing and physical-distancing rules, and so is the COVID-19 vaccine acceptance. Governance and pandemic preparedness reflect the national capacity for effective pandemic response strategies and influence the public’s support for these strategies. Building greater public sector competence and consequent trust in government is a priority for policymakers to promote public compliance with pandemic mitigation strategies including vaccination.

Culture of uncertainty avoidance explained 25.8% of cross-country variation in COVID-19 vaccine refusals expressed on Twitter. This is consistent with a previous study on the effect of social values on vaccine acceptance.^26^ Increased vaccine hesitancy and refusal associated with conservative political views are attributed, in part, to concerns about vaccine safety and effectiveness.^27^ People usually have less risk tolerance for vaccines than medicine due to usage for healthy individuals and the invisibility of disease threats. A culture of avoiding uncertainty and prevalent misinformation amplified the perceived risk of vaccination and led to vaccine refusal, especially for the newly developed COVID-19 vaccine. It is necessary to provide accurate, timely information about the vaccine and clearly communicate the relevant risk. Early responses to vaccine misinformation are also needed. These would relieve the public’s concerns about vaccine safety and boost their trust in vaccines.

Moreover, our study highlighted continuously declining trends of COVID-19 vaccine acceptance and confidence with a temporary uptrend following its rollout worldwide. A similar global declining trend in positive attitudes toward COVID-19 vaccination was also observed in a cross-country survey, with a decline of approximately 8% from February to July 2021.^28^ Canadian tweets between December 2020 and May 2021 also showed that positive sentiment for vaccination declined since March 2021.^29^ The declining support from the public posed a global challenge for policymakers to rely on vaccination to end the pandemic.^28,30^ The government should prepare early to maintain public support for vaccination.

Our study further provides evidence that increased online searches for AEFI were a marker of a global decline in COVID-19 vaccine acceptance. Similarly, two Canadian studies also considered severe AEFI concerns to be a major cause of the global decline in COVID-19 vaccination acceptance and confidence.^28,29^ The safety of COVID-19 vaccines was of great concern to the public due to the rapid timeline of vaccine development, fears of new, innovative vaccine development technologies such as mRNA, and reports on AEFIs. As mass COVID-19 vaccination progressed, many countries reported severe, if rare, AEFIs such as thrombosis, anaphylaxis, myocarditis, TAFRO syndrome, and Guillain-Barre syndrome since March 2021, accompanied by a surge in Google® search volume on AEFIs and a decreasing trend in COVID-19 vaccine acceptance globally.^31-33^

COVID-19 vaccines are among the safest vaccines ever developed with a low frequency of severe AEFIs (< 0.1%),^31^ yet the massive exposure and dissemination of AEFI-related news fueled public anxiety about COVID-19 vaccines and triggered a high volume of web searches. This high online AEFI-related search volume reflects widespread concern for AEFIs. As a mediator influencing individual attitudes toward COVID-19 vaccination, it can be the target of informative surveillance using social media.^34^

Our study’s strengths include the large number of countries and territories sampled, the number of languages examined, and the apparent success of our machine learning approaches. Consistency between social media tweets and surveys on COVID-19 vaccine acceptance and its correlation with vaccine coverage verified the reliability of social media surveillance. The study’s limitations are as follows. First, COVID-19 vaccination-related opinions on Twitter® evolve and vary; real-time monitoring of vaccine acceptance is needed in the future to provide more updated information, rather than a retrospective view. We believe that our strategy can be modified towards real-time use. Second, this study established correlations, but ecological studies do not prove causality. Finally, the performance of the deep learning model could vary across languages; this is a limitation shared by all multilingual deep learning studies and is the topic of ongoing refinement of deep learning algorithms.

In summary, social media surveillance using machine learning can address complex public health issues such as vaccine acceptance or hesitancy in many venues and in many languages. We believe this to be a new frontier for public health and medical surveillance, permitting rapid input to agencies and policymakers as to public perceptions and viewpoints. Recognition of fears and their origins is the first step in rapid educational response and anticipation of analogous views in future similar situations. In future pandemics, newly-developed vaccines may face the potential low-level and declining trend in acceptance, like COVID-19 vaccines, and government should prepare early to maintain public support for vaccination.

## Supporting information

Supplementary Material 1_Results

## Data Availability

All data described in the results and Python/R codes for data analysis/visualization are shared on GitHub, upon acceptance of this paper. Original tweets are not shared according to Twitter's data policy. Other data and codes are available on request to the corresponding author.

https://github.com/xinyuuzhou/COVID-19-vaccine-on-Twitter

## Ethical statement

The study was approved by the Institutional Review Board of the School of Public Health, Fudan University (IRB#2022-01-0938).

## Contributors

ZH and X. Zhou co-conceived and designed the study. LL, HJL, and X. Zhou acquired Twitter data. X. Zhang and SZ collected data other than Twitter. X. Zhou trained and deployed deep learning models, implemented statistical analyses, and created the figures. ZH and X. Zhang verified the data. X. Zhou, ZH, and X. Zhang wrote the manuscript. ZH and LL supervised the study. All authors had access to the data, contributed to data interpretation, and critically revised and approved the manuscript.

## Data sharing statement

All data described in the results and Python/R codes for data analysis/visualization are shared on <u>GitHub</u>, upon acceptance of this paper.Original tweets are not shared according to Twitter’s data policy. Other data and codes are available on request to the corresponding author.

## Acknowledgements

Zhiyuan Hou acknowledges financial support from the National Natural Science Foundation of China (71874034), and the Soft Science Research Project of Shanghai “Science and Technology Innovation Action Plan” (22692107600). We thank the following students from the School of Public Health, Fudan University: Ying Zhang for her help with data collection; Yixing Tong, Fanxing Du, Linyao Lu, Sihong Zhao, and Kexin Yu for their help on data annotation; and Bo Zheng for his help on data analysis. We also thank Chen Wang from the Software School of Fudan University for his helpful suggestions on fine-tuning deep learning models.

