## Supplementary Material 1_Methods for "Global spatiotemporal trends and determinants of COVID-19 vaccine acceptance on Twitter: a multilingual deep learning study in 135 countries and territories"

**Supplementary Material I** for the manuscript "Global spatiotemporal trends and determinants of COVID-19 vaccine acceptance on Twitter: a multilingual deep learning study in 135 countries and territories."

#### Section I - Supplementary Methods

### Supplementary Methods:

This section provides a detailed description of our methods.

#### 1. Multilingual data curation

**Identify multilingual keywords**

To collect multilingual data on Twitter® concerning the COVID-19 vaccine, the first step is to identify all relevant keywords. In this step, we collected, expended, and verified keywords collected by professional translators from a large translation company in China (Beijing Chinese-Foreign Translation & Information Service Co., Ltd., "译鱼人工翻译", http://www.cipgtrans.com/).

Firstly, we instructed the company to identify translators for 97 languages (the 100 languages supported by XLM-RoBERTa (XLM-R) model except for the three languages the authors know: English, Traditional Chinese, and Simplified Chinese). The company identified translators in 87 languages. We then asked the translators to write down all keywords relevant to the COVID-19 vaccine in the language they specialise in. Sample keywords and detailed instructions for identifying keywords were provided to the translators in Chinese and English, and they identified 483 keywords in 87 languages. Meanwhile, the researchers identified 14 keywords in English, Simplified Chinese, and Traditional Chinese. As a result, we collected a total of 497 keywords in 90 languages.

In most languages, "COVID", "COVID-19", and "COVID19" all mean the same. However, the translators might believe only one or two of them are the keywords and neglect the rest of them[1]. Therefore, if a keyword contains any of "COVID-19", "COVID19", or "COVID" in the original list of manually translated keywords, researchers will ensure the remaining one/ones in the keyword list. This way, we expanded the list of manually translated keywords (to 709 keywords).

---

[1] For example, "Vaksinimi kundër COVID" is in the preliminary list of human-translated Albania keywords, while "Vaksinimi kundër COVID19" is not.

To further supplement the list of keywords, using Google® Translate[2], we collected multilingual machine translations of nine selected English keywords concerning COVID-19 vaccine (“COVID vaccine”, “COVID vaccines”, “COVID vaccination”, “COVID-19 vaccine”, “COVID-19 vaccines”, “COVID-19 vaccination”, “coronavirus vaccine”, “coronavirus vaccines”, and “coronavirus vaccination”)[3]. Duplicated keywords were removed, and our list expended to 1062 keywords.

Finally, our team manually checked and ensured all keywords collected in earlier steps were COVID-19 vaccine-related. We used Google® Translate to check the keywords’ machine translations in English and Chinese. Keywords that were not translated into COVID-19 vaccine/vaccination (or their equivalents) in both English and Chinese were removed. We then used Google® Image Search to verify the remaining keywords. Keywords whose first page of search results contained more than two images irrelevant to the COVID-19 vaccine were removed. In the end, we identified all eligible keywords in 90 languages (n=1027) (keywords are available in supplementary material 2).

An overview of the keyword selection strategy can be found in Figure S1.

**Figure S1.** Keyword selection strategy

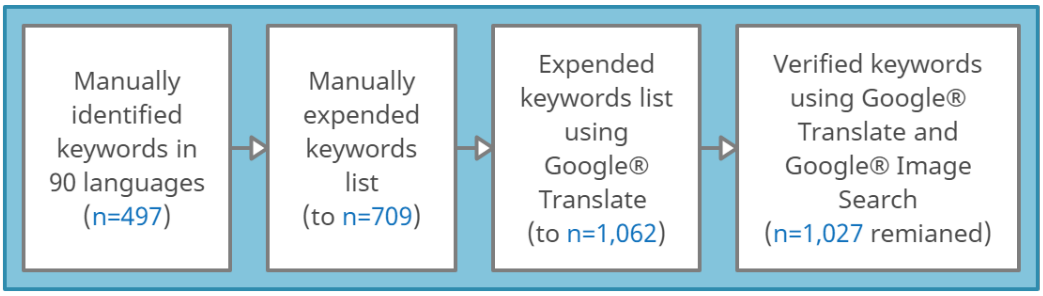

**Collection of data from Meltwater®**

Using the multilingual keywords, we collected 13,093,406 multilingual tweets on Twitter® in the public domain regarding COVID-19 vaccination across the globe from November 13, 2020, to

---

2 https://translate.google.com/

3 We collected machine translations in 86 languages supported by Google Translate. One of the languages (Burmese) was not supported by Google Translate (in February 2022).

March 5, 2022, through Meltwater® Social Listening Tool[4].

During data collection, tweets that contain Twitter® users' own comments, including original tweets and quotes, were collected. On the other hand, retweets didn't allow users to comment and were not collected[5]. The difference between retweets and quotes is further illustrated in Twitter® Help Centre[6]. During data collection, tweets with the same content sent by the same user were removed.

#### 2. Manually annotating English-language Tweets

To leverage deep learning models for tweet annotation, a common practice is to annotate tweets by humans and ask the model to imitate human annotation. Our team developed an annotation framework for COVID-19 vaccine-related tweets based on the framework of vaccine hesitancy proposed by the WHO.(SAGE; manual annotation study) We validated the framework using a 500-tweet sample. Based on the framework, the research team manually annotated 8,125 English-language tweets on COVID-19 vaccination into eight predefined categories. The definition of each category is available in Table S2. Each tweet was annotated by two annotators independently, and a third annotator resolved disagreements.

Tweets were annotated in two steps. Firstly, annotators determine whether each tweet was sent by human users. Tweets from bot accounts, news reports, advertisements, and governmental announcements, were removed and archived. Second, the human-generated tweet would be annotated one by one according to their relevance to the eight categories in our framework, including two categories concerning COVID-19 vaccine acceptance: (1) intent to accept COVID-19 vaccination, (2) intent to reject COVID-19 vaccination, and six determinants of acceptance: (3) belief that COVID-19 vaccine is effective, (4) belief that COVID-19 vaccine is not safe, (5) misinformation or rumours on COVID-19 vaccine, (6) distrust in government, (7) (perceived) COVID-19 vaccine accessibility, (8) (perceived) COVID-19 vaccine distribution. Depending on the tweet content, tweets could be annotated into one category, multiple categories, or no category.

---

4 https://www.meltwater.com/
5 For the same reason, we dropped all quotes without user's comment.
6 https://help.twitter.com/en/using-twitter/types-of-tweets

#### 3. Finetune multilingual deep learning models using manually annotated tweets

**About the XLM-R model**

XLM-RoBERTa (XLM-R) is a cutting-edge pretrained transformers-based deep neural network for analysing multilingual textual data. In diversified benchmarks for multilingual models, it significantly outperformed other widely-used state-of-the-art multilingual models, such as XLM and multilingual BERT, especially in low-resource languages, such as Urdu and Swahili [(XLM-R)]. In these benchmarks, multilingual models' performance was evaluated in the multi-lingual dataset, but they were finetuned purely with mono-lingual manually annotated datasets. XLM-R is pre-trained using 2.5 TB of filtered CommonCrawl data in the 100 most spoken languages. The model was pre-trained using the Masked Language Models objective in a self-supervised style. During the pre-training process of the model, language materials in each of the 100 languages were provided to the model, and some parts (15%) of the text were randomly masked. The model is asked to predict the masked words as correctly as possible, and after massive computation, it comprehends 100 languages simultaneously. As a result, once the pre-trained model is fine-tuned with a small, manually annotated monolingual downstream training data, it can precisely analyse multilingual data in around 100 languages. In the XNLI test, the XLM-R model finetuned with English-language data reached 80.9% accuracy in cross-lingual transfer, outperforming XLM by 10.2% and multilingual BERT by 14.6%; it not only got high accuracy in high-resource languages (English and French at 89.1% and 84.1%, respectively) but also performed well in low-resource languages (Swahili and Urdu at 73.9% and 73.8%, respectively). In this study, we employed the 24-layer version of the pre-trained XLM-R model, which contains 550 million parameters.

**Data augmentation and hyperparameters**

To deploy the model for analysing tweets on the COVID-19 vaccine, the pre-trained XLM-R model needed to be finetuned using our manually annotated COVID-19 tweets dataset. We randomly selected 80% of our annotated tweets as the training set, 10% as the validation, and 10% as the test set. Our manually annotated data was unbalanced. Before finetuning the model, we employed data augmentation to enlarge the number of positive labels in training set to enhance the model's performance. We hired two approaches for data augmentation: 1) back translation using Baidu Translate API (http://api.fanyi.baidu.com/), and 2) simulating spelling errors using the python package "nlpaug". We chose the models' hyperparameters based on the model performance in our validation set. The hyperparameters are available in Table S1.

**Table S1**. Settings to fine-tune the deep learning models.

| XLM-RoBERTa Model (a): find human-generated posts, N(training set)=**6581**, N(validation set)=**732** | | | | | | |
|---|---|---|---|---|---|---|
| Category | n(labelled) before data augmentation | n(labelled) after data augmentation | batch size | learning rate | epoch | Data augmentation |
| Human-generated | 3753 | 3753 | 16 | 2.00E-05 | 4 | no |
| XLM-RoBERTa Model (b): classify, N(original training set)=**3377**, N(validation set)=**376** | | | | | | |
| Category | n(labelled) before data augmentation | n(labelled) after data augmentation | batch size | learning rate | epoch | Data augmentation |
| Vaccination intent | | | | | | |
| (a) Intent to accept COVID-19 vaccination | 1911 (1720+191) | 1911 (1720+191) | 32 | 2.00E-05 | 4 | no |
| (b) Intent to reject COVID-19 vaccination | 489 (440+49) | 1369 (1320+49) | 24 | 2.00E-05 | 3 | yes |
| Vaccine confidence | | | | | | |
| (c) Belief that COVID-19 vaccine is effective | 671 (604+67) | 1275 (1208+67) | 16 | 1.00E-05 | 4 | yes |
| (d) Belief that COVID-19 vaccine is not safe | 300 (270+30) | 570 (270+30) | 32 | 2.00E-05 | 3 | yes |
| Information environment | | | | | | |
| (e) Misinformation or rumours on COVID-19 vaccine | 345 (310+35) | 1275 (1240+35) | 32 | 2.00E-05 | 3 | yes |
| (f) Distrust in government | 445 (400+45) | 1245 (1200+45) | 16 | 2.00E-05 | 4 | yes |
| Vaccine convenience | | | | | | |
| (g) COVID-19 vaccine accessibility | 297 (267+30) | 831 (801+30) | 16 | 2.50E-05 | 3 | yes |
| (h) COVID-19 vaccine equity | 587 (528+59) | 1115 (1056+59) | 24 | 2.00E-05 | 4 | yes |

**Measuring the performance of machine learning: $F_1 - score$ and $accuracy$**

$F_1 - score$ and $accuracy$ are widely used in Natural Language Processing to measure the performance of machine learning models. The models are asked to predict the human annotations in the test set, and we compare the machine's annotation with the human's. Here, human annotation is considered the ground truth, a common practice in machine learning. Like other studies, $F_1 - score$ and $accuracy$ are calculated as follows.

$$F_1 - score = 2 \cdot \frac{1}{\frac{1}{accuracy} + \frac{1}{recall}} = 2 \cdot \frac{accuracy \cdot recall}{accuracy + recall}$$

Where,

$$accuracy = \frac{True\ Positive}{True\ Positive + False\ Positive}$$

$$recall = \frac{True\ Positive}{True\ Positive + False\ Negative}$$

In the test set, we evaluated the model performance. As shown in Table S2, the $accuracy$ of models used in this study ranges from 72.73% to 89.74%, and the $F_1 - score$ ranged from 73.0% to 86.6%. The models were trained, validated, and tested on AutoDL (https://autodl.com/) (Python 3.8).

**Table S2.** Definitions of the categories and the performance of deep learning.

| **Finetuned XLM-RoBERTa Model (a): to find human-generated posts, N(test set)=812** | | | | |
|---|---|---|---|---|
| Category | Definition | n | $F_1$-score | Accuracy |
| Human-generated | Tweets sent by human users on COVID-19 vaccine. (Ads, news, tweets from bot accounts or tweets irrelevant to COVID-19 vaccine were removed.) | 435 | 0.8662 | 0.8782 |
| **Finetuned XLM-RoBERTa Model (b): to classify, N(test set)=435** | | | | |
| Category | Definition | n | $F_1$-score | Accuracy |
| Vaccination intent | | | | |
| (a) Intent to accept COVID-19 vaccination | Twitter® posts indicating that they will accept, support or be willing to get COVID-19 vaccination. | 222 | 0.8541 | 0.8964 |
| (b) Intent to reject COVID-19 vaccination | Twitter® posts indicating that they will reject, do not support, or be unwilling to get COVID-19 vaccination. | 54 | 0.7304 | 0.7778 |

| | | | | |
|---|---|---|---|---|
| Vaccine confidence | | | | |
| (c) Belief that COVID-19 vaccine is effective | Twitter® users had confidence in the effectiveness of the COVID-19 vaccine, believing that it is effective. | 82 | 0.8098 | 0.8049 |
| (d) Belief that COVID-19 vaccine is not safe | Twitter® users lacked confidence in the safety of the COVID-19 vaccine, believing it was not safe. | 34 | 0.6753 | 0.7647 |
| Information environment | | | | |
| (e) Misinformation or rumours on COVID-19 vaccine | Negative information about all vaccines on tweets, such as misinformation, rumours, anti-vaccine campaigns, anti-intellectual, anti-science campaigns, and vaccine scandals. | 34 | 0.7500 | 0.6176 |
| (f) Distrust in government | Twitter® users indicated distrust in government or policy-makers, including all-level government, ministry of health, CDC, etc. | 55 | 0.7921 | 0.7273 |
| Vaccine access | | | | |
| (g) COVID-19 vaccine accessibility | Tweets mentioned the production or supply capacity of the COVID-19 vaccine or the self-efficacy of accessing it. | 41 | 0.6818 | 0.7317 |
| (h) COVID-19 vaccine equity | Tweets mentioned (priority) vaccination groups or vaccine allocation equity | 64 | 0.8088 | 0.8594 |
| **micro average** | | | **0.7973** | **0.8174** |

#### 4. Deep learning-based annotation of tweets

An overview of the data collection and analysis process is available in Figure 1. This study fine-tuned XLM-R deep learning model to annotate tweets like human annotators. 13,093,406 unique tweets were collected, and similar to the manual annotation process, we first identified if tweets were sent by humans or not. Tweets deemed not to be sent by a human (n=6,046,183) were removed, and tweets sent by humans (n=7,047,223) were further classified using eight fine-tuned XLM-RoBERTa binary classifiers to identify if the human-generated tweets expressed any of the following categories, including:

Two categories concerning COVID-19 vaccine acceptance:

- "Intent to accept COVID-19 vaccination"
- "Intent to reject COVID-19 vaccination"

Six categories concerning the determinants of COVID-19 vaccine acceptance:

- "Belief that COVID-19 vaccine is effective"
- "Belief that COVID-19 vaccine is not safe"
- "Misinformation or rumours on COVID-19 vaccine"
- "Distrust in government"
- "(perceived) COVID-19 vaccine accessibility"
- "(perceived) COVID-19 vaccine distribution"

The definitions of the categories are available in Table S2.

#### 5. Triangulating data & statistical analysis

Tweets are aggregated according to their metadata regarding time and geo-location provided by Meltwater®. We calculated the opinion of each Twitter user by averaging all his/her tweets within each time interval. We then calculate the average opinions of Twitter® users spatiotemporally.

**Spatial trends**

Spatial trends of COVID-19 vaccine acceptance and its determinants on Twitter were estimated at the country level. Among all 7,047,223 human-generated tweets, 4,137,550 (58.7%) tweets were geo-located. We included countries and territories meeting the following criteria: (1) Twitter® is not banned by the relevant government, (2) there are data from at least 100 Twitter® users in that country or territory, (3) the analyzed 90 languages cover all official languages in that country or territory. There were 171 countries and territories with tweets sent by at least 100 Twitter® users, and their censorship status and official languages are shown in Supplement table #2. After excluding those with official languages not covered by our dataset and those banned Twitter®, we included 135 countries or territories.

**Determinants of spatial trends**

To explore the determinants of the spatial trends in COVID-19 vaccine acceptance on Twitter®, we first calculated their correlation to vaccine confidence, information environment, and perceived vaccine convenience on Twitter®. We further collected 20 country-level indicators regarding governance, pandemic preparedness, trust, culture, social development status, and demographics from multiple external sources (supplementary material Table S4) and evaluated their correlations to COVID-19 vaccine acceptance on Twitter using univariant linear regression.

**Linkages to COVID-19 vaccine uptake**

To explore the public health implication of COVID-19 vaccine acceptance on Twitter, we linked country-level acceptance to country-level COVID-19 vaccine coverage using univariant linear regression. In the supplementary material, we further explored the linkages using multilevel regression.

**Temporal trends**

Global temporal trends are calculated at the daily level. Spline regression was employed to fit global temporal trends in opinions of the COVID-19 vaccine on Twitter. Country-level trends were calculated at the weekly and monthly levels in countries with sufficient data.

**Linkages to temporal trend**

We obtained weekly country-level Google® Search Trends data on 12 topics related to adverse events following immunisation (AEFI). The 12 topics were selected based on our team's knowledge about AEFI and verified using the five most relevant topics and queries according to Google® Trends (Supplement Worksheet #4). We utilized univariant linear regression to explore their associations to weekly-level trends of COVID-19 vaccine acceptance on Twitter in each country.

### External data utilised in this study

##### Table S4. Country-level indicators used in association analyses

| Indicators | Description | Data source | Time range | Value range |
|---|---|---|---|---|
| **COVID-19 vaccination coverage** | | | | |
| **COVID-19 vaccination coverage, %** | The total number of doses divided by the total population of the country. | Our World in Data | Nov 13, 2020 – Mar 5, 2022 | .48-121.53 |
| **Governance** | | | | |
| **Government effectiveness** | Perceptions of the quality of public services, the quality of the civil service and the degree of its independence from political pressures, the quality of policy formulation and implementation, and the credibility of the government's commitment to such policies. | World Bank | 2020 | -2.31-2.34 |
| **Control of corruption** | Perceptions of the extent to which public power is exercised for private gain, including petty and grand forms of corruption, and "capture" of the state by elites and personal interests. | World Bank | 2020 | -1.71-2.27 |
| **State fragility** | Indicator P2: Incapacity to provide essential public goods and services and cope with shocks | Fragility States Index | 2022 | .90-10 |
| **Pandemic preparedness** | | | | |
| **Epidemic Ready Score** | The ready score determines whether a country is prepared to find, stop, and prevent epidemics using data from the WHO's Joint External Evaluation. There are 19 areas of preparedness and response capacity scored. | Prevent Epidemics | Latest | 26-93 |
| **Global Health Security Index** | The GHS index includes six categories: prevention, detection and reporting, rapid response, health systems, compliance with international norms, and risk environment. Although the GHS Index can identify preparedness resources and capacities available in a country, it cannot predict whether or how well a country will use them in a crisis. | GHS Index | 2021 | 16-75.90 |
| **Doctors per 1000 people** | Health systems resources | The global health observatory indicators | Latest | .23-84.20 |
| **Trust** | | | | |
| **Trust in government, %** | Trust coded as "A lot" or "Some" on W5B asking about confidence in government | Wellcome Global Monitor 2020 COVID-19 | 2017-2021 | 16.14-93.70 |

| | | | | |
|---|---|---|---|---|
| **Trust in science, %** | Trust coded as "A lot" or "Some" on W6 asking about trust in science | Wellcome Global Monitor 2020 COVID-19 | 2018 | 47.17-96.70 |
| **Interpersonal trust, %** | Trust coded as "most people can be trusted" on Q57 asking about interpersonal trust | World values survey wave 7 | 2021 | 4.25-56.58 |
| **Culture-related index** | | | | |
| **Individualism** | The ties between individuals are loose: everyone is expected to look after him/herself and his/her immediate family | Clearly Cultural | Latest | 6-91 |
| **Uncertainty avoidance** | Uncertainty avoidance deals with a society's tolerance for uncertainty and ambiguity. It indicates to what extent a culture programs its members to feel either uncomfortable or comfortable in unstructured situations (novel, unknown, surprising, and different from usual). | Clearly Cultural | Latest | 8-112 |
| **Social development status** | | | | |
| **Socio-Demographic Index** | The Socio-demographic Index (SDI) is a composite indicator of development status strongly correlated with health outcomes. It is the geometric mean of 0 to 1 indices of total fertility rate under the age of 25 (TFU25), mean education for those ages 15 and older (EDU15+), and lag distributed income (LDI) per capita. | Global Burden of Disease | 2019 | .08-0.93 |
| **Ln (GDP per capita)** | Natural logarithm of GDP per capita (constant US＄) | World Bank | 2021 | 6.10-11.82 |
| **School enrollment, %** | The ratio of total enrollment, regardless of age, to the population of the age group. | World Bank | 2020 | 1-149 |
| **Internet coverage, %** | Proportion of individuals using the Internet in total population. The Internet can be used via a computer, mobile phone, personal digital assistant, games machine, digital TV etc. | World Bank | 2020 | 10-100 |
| **Demographics** | | | | |
| **Population density** | People per sq. km of land area | World Bank | 2020 | 2-8019 |
| **Population ages 0-14, %** | Percentage of population age 0-14 in total population | World Bank | 2021 | 12.27-46.73 |
| **Population ages 65 and above, %** | Percentage of population ages 65 and above in total population | World Bank | 2021 | 1.45-28.70 |
| **Urban population, %** | Percentage of urban population in total population | World Bank | 2021 | 18.86-100 |
