## Supplementary Material 1_Results for "Global spatiotemporal trends and determinants of COVID-19 vaccine acceptance on Twitter: a multilingual deep learning study in 135 countries and territories"

**Supplementary results for the manuscript**
"Global spatiotemporal trends and determinants of COVID-19 vaccine acceptance on Twitter: a multilingual deep learning study in 135 countries and territories."

**Supplementary material I section II: Supplementary Results**

### Section II: Supplementary Results

**Table S5.** Twitter users count, sorted by the number of tweets.

| *number of tweets* | *number of users* | *percentage* |
|---|---|---|
| *1* | 2320786 | 69.40% |
| *2* | 486,166 | 14.54% |
| *3* | 192,091 | 5.74% |
| *4* | 100,228 | 3.00% |
| *5* | 59,471 | 1.78% |
| *6* | 39,208 | 1.17% |
| *7* | 27,188 | 0.81% |
| *8* | 20,478 | 0.61% |
| *9* | 15,341 | 0.46% |
| *10* | 11,710 | 0.35% |
| *11-20* | 47,437 | 1.42% |
| *21-30* | 12,368 | 0.37% |
| *31-40* | 4,908 | 0.15% |
| *41-50* | 2,460 | 0.07% |
| *51-100* | 3,450 | 0.10% |
| *⩾101* | 854 | 0.03% |
| *sum* | 3,344,144 | 100.00% |

The table above illustrates how many human-generated tweets were sent by each user. Overall, 7,047,223 tweets analysed in this study were sent by a total of 3,344,144 users. The majority of the users ($69.40\%, N_{\text{user}} = 2{,}320{,}786$) only sent one tweet and most users ($97.86\%, N_{\text{user}} = 3{,}272{,}66$) sent less than 10 tweets. However, there are still 854 users who sent over 100 tweets.

**Figure S2.** Spatiotemporal trends of tweet count.

**Figure S2a.** Daily number of human-generated tweet.

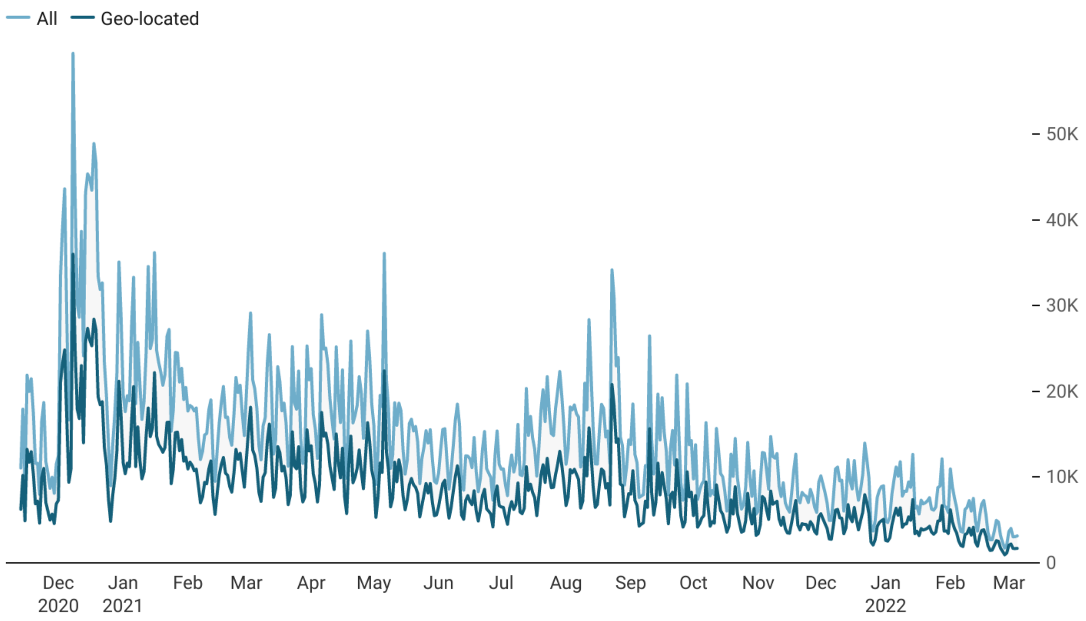

**Figure S2b.** Weekly proportion of tweet count at the country-level.

- Each point in the figure is calculated using the following formula.

$$Proportion = \frac{\text{The number of tweets sent by human in a specific week (in the country/territory)}}{\text{The number of tweets sent by human from November 13,2020 – March 5,2022 (in the country/territory)}}$$

- A point with a deeper colour suggests a higher percentage of tweets are sent during the period.

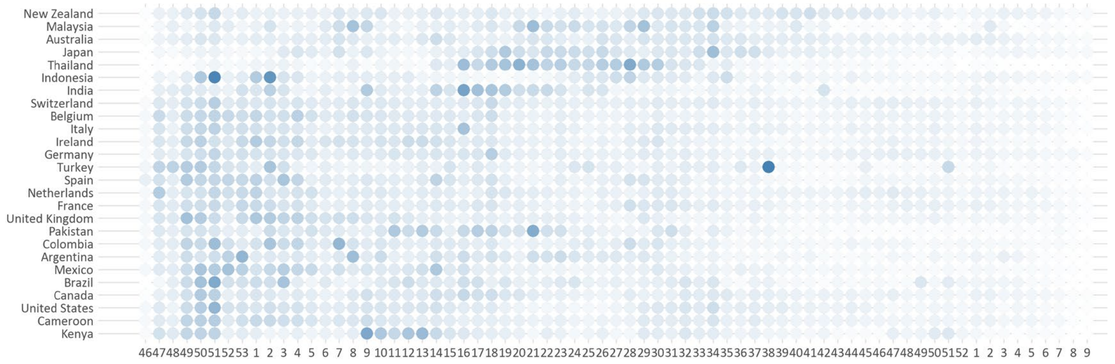

**Figure S3.** Regional variations in COVID-19 vaccine acceptance, vaccine confidence, information environment, and prevalence of vaccine access-related discourse among Twitter users, November 13, 2020 – March 5, 2022.

- Each point in the plot represents a country or territory.

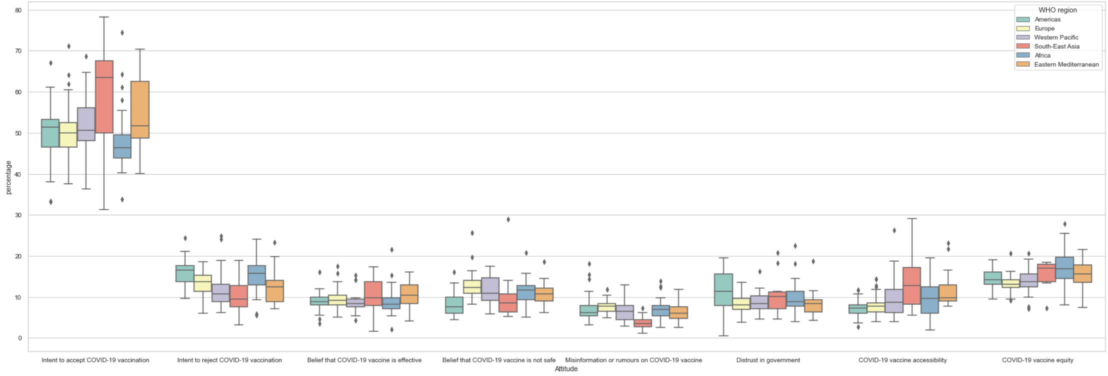

**Figure S4.** Multilevel regression coefficients between COVID-19 vaccine acceptance on Twitter and COVID-19 vaccine coverage in each month.

- Countries and territories are grouped according to WHO regions.

**Figure S4a.** COVID-19 vaccine acceptance v.s. monthly coverage

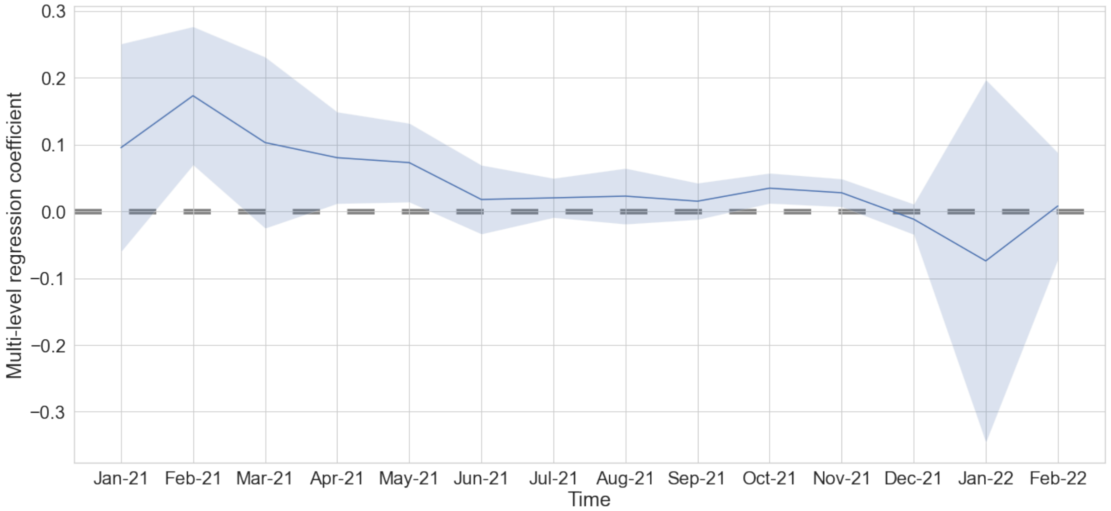

**Figure S4b.** COVID-19 vaccine rejection v.s. monthly coverage

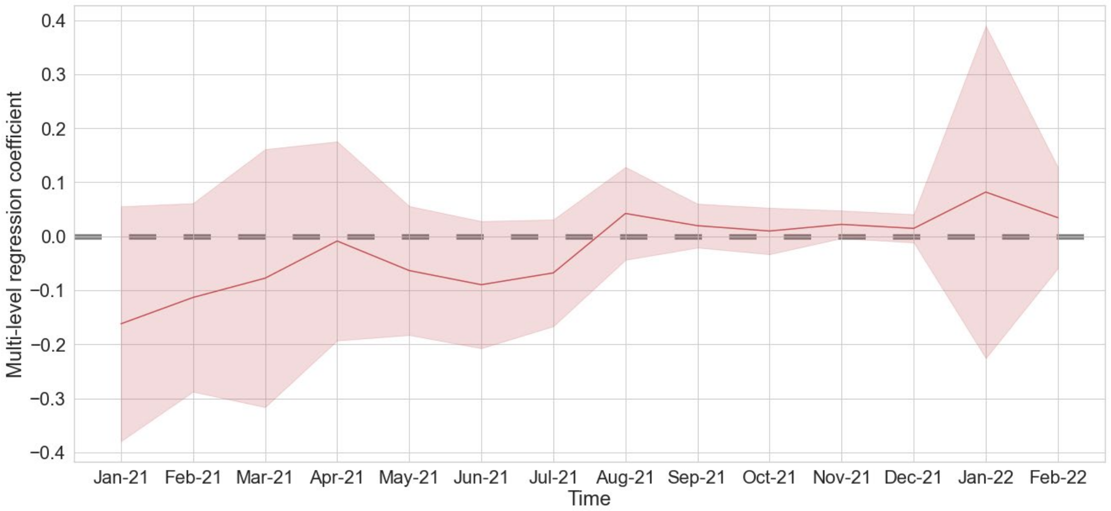

**Table S6.** Association between Twitter users' acceptance and coverage of COVID-19 vaccination across 131 countries and territories

**Table S6a.** Results from univariant linear regressions.

| Variables | COVID-19 vaccine coverage | | | |
|---|---|---|---|---|
| | **Coefficient** | **95% CI[a]** | **p-value** | **$R^2$** |
| **Intent to accept COVID-19 vaccination** | ***1.007*** | ***[0.389,1.625]*** | ***0.002*** | ***7.5%*** |
| **Intent to reject COVID-19 vaccination** | ***-1.983*** | ***[-3.166,-0.800]*** | ***0.001*** | ***7.9%*** |

[a] Confidence Interval.

**Table S6b.** Results from multi-level regression analyses.

| Variables | COVID-19 vaccine coverage | | | |
|---|---|---|---|---|
| | **Coefficient** | **Std.Error** | **z value** | **Pr(>\|z\|)** |
| **Multi-level regression, grouped by WHO regions** | | | | |
| **Intent to accept COVID-19 vaccination** | ***0.782*** | ***0.242*** | ***3.226*** | ***0.001*** |
| **Intent to reject COVID-19 vaccination** | ***-1.223*** | ***0.475*** | ***-2.576*** | ***0.010*** |
| **Multi-level regression, grouped by World Bank's national income levels** | | | | |
| **Intent to accept COVID-19 vaccination** | ***0.929*** | ***0.208*** | ***4.466*** | ***0.000*** |
| **Intent to reject COVID-19 vaccination** | ***-1.111*** | ***0.430*** | ***-2.582*** | ***0.010*** |

**Figure S5.** Regional weekly-level trends of COVID-19 vaccine acceptance, vaccine confidence, information environment, and prevalence of vaccine access-related discourse among Twitter users, November 13, 2020 – March 5, 2022

- Each point in the figure represents the weekly-level prevalence of a specific category among all Twitter users in a given region.
- A point with a deeper colour suggests that a higher proporton of Twitter users expressed the specific opinion.

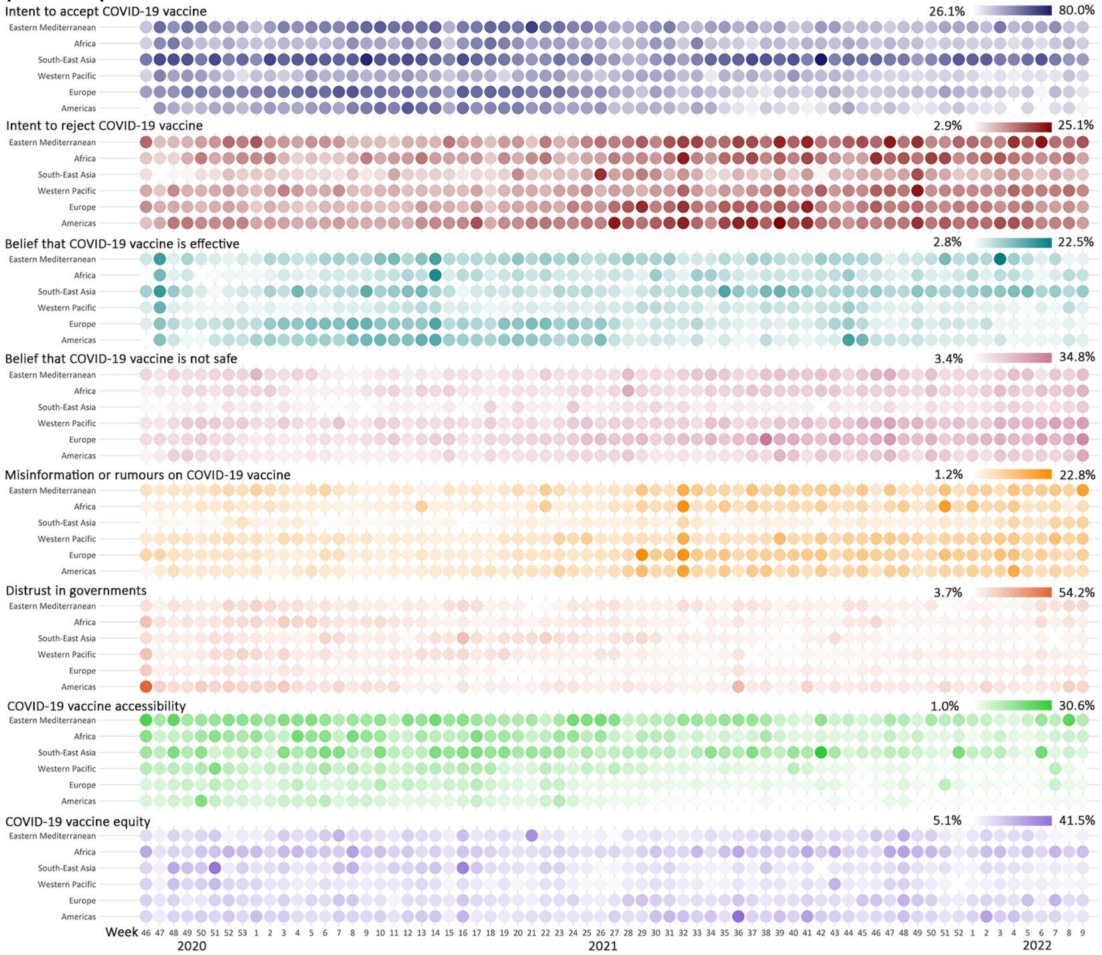

**Figure S6.** Weekly-level trends of weighted COVID-19 vaccine acceptance among Twitter users in 26 countries, November 13, 2020 – March 5, 2022

- Only the 26 countries or territories with sufficient Twitter data (total number of Twitter users mentioning COVID-19 vaccination >5000; weekly number >28) were shown.

**Figure S6a.** Weekly-level trends of weighted COVID-19 vaccine acceptance.

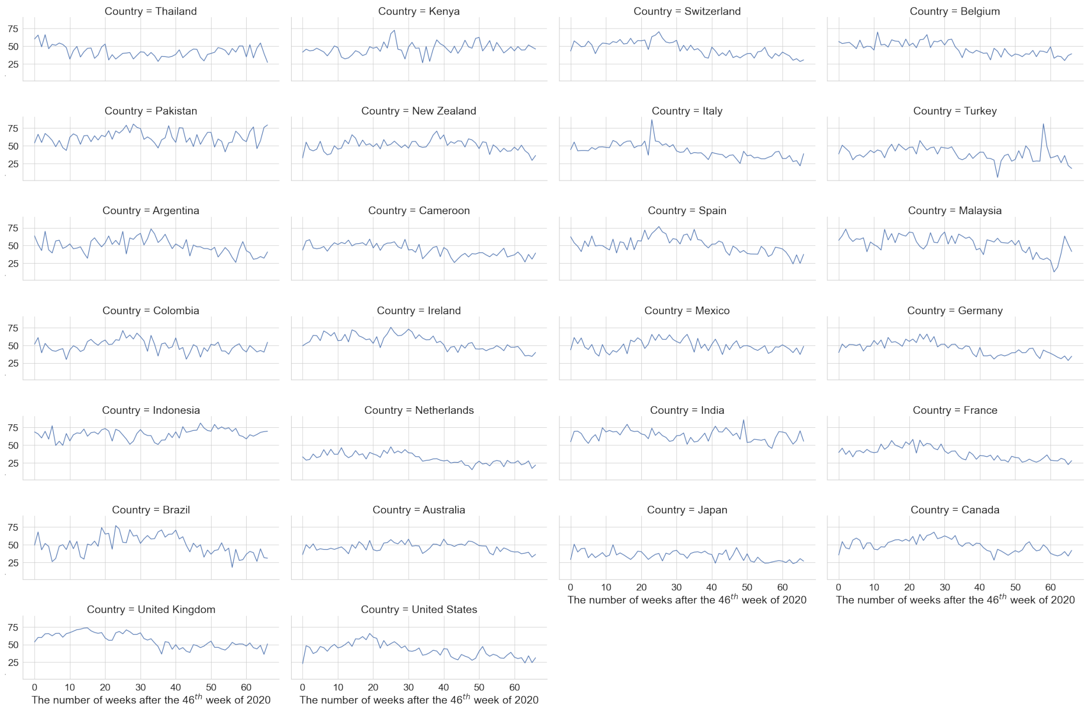

**Figure S6b.** Weekly-level trends of weighted COVID-19 vaccine refusal.

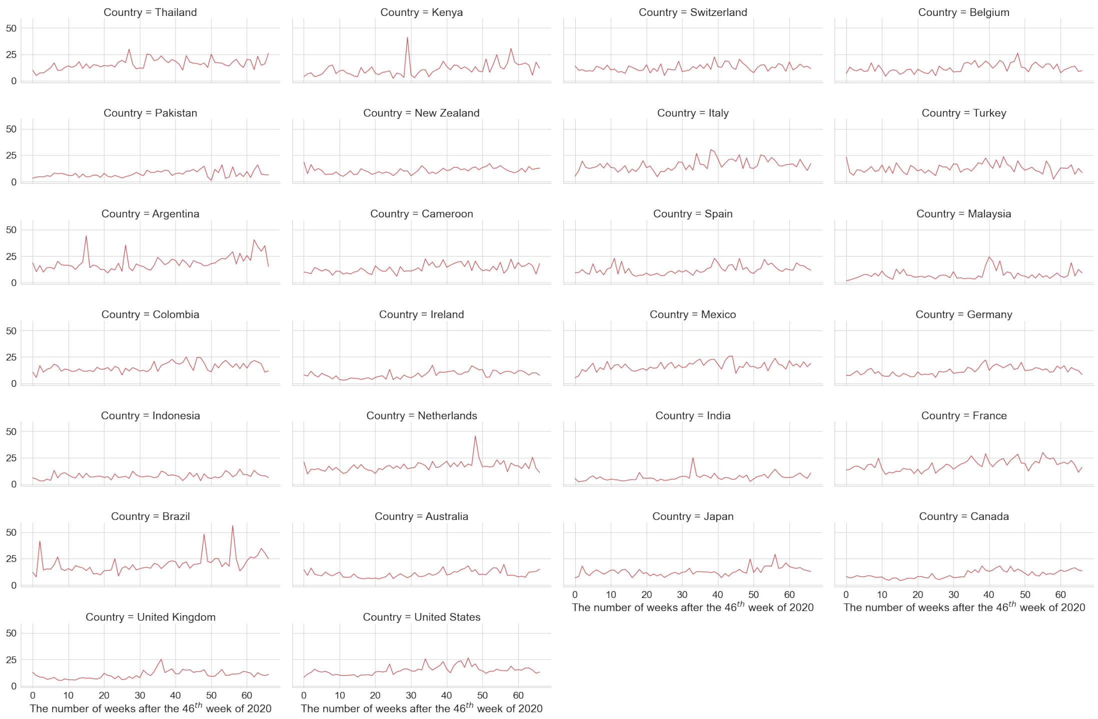

**Figure S7.** Global trends on vaccine adverse event, January 2004 - February 2022

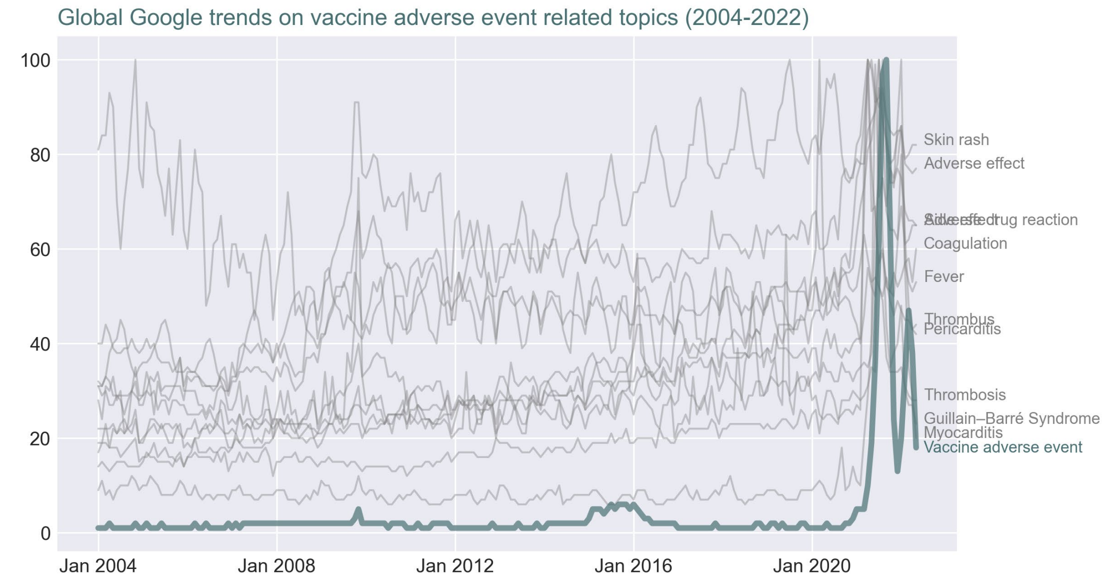

**Figure S8.** Weekly trends of 12 AEFI-related Google Trends topics in 26 countries or territories, November 2020 – February 2022

**Adverse drug reaction**

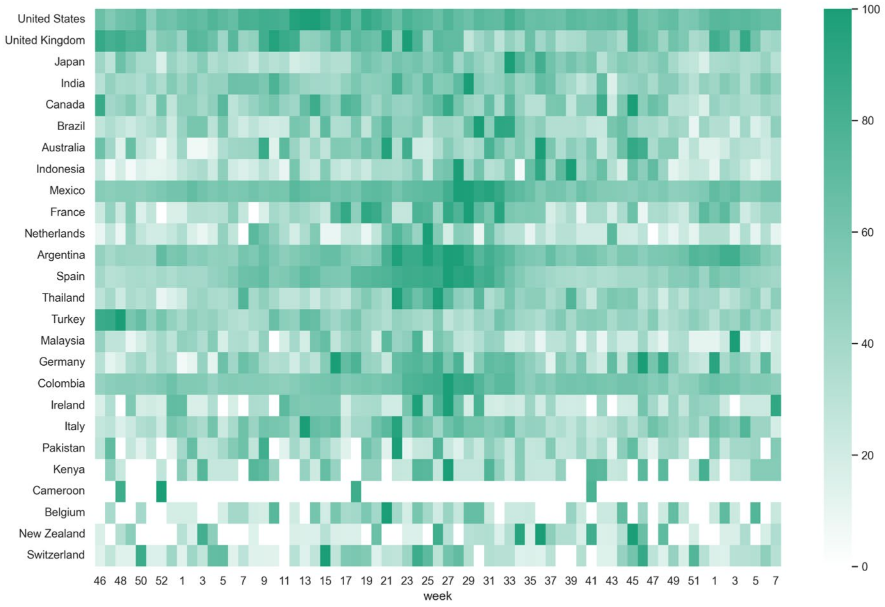

**Adverse effect**

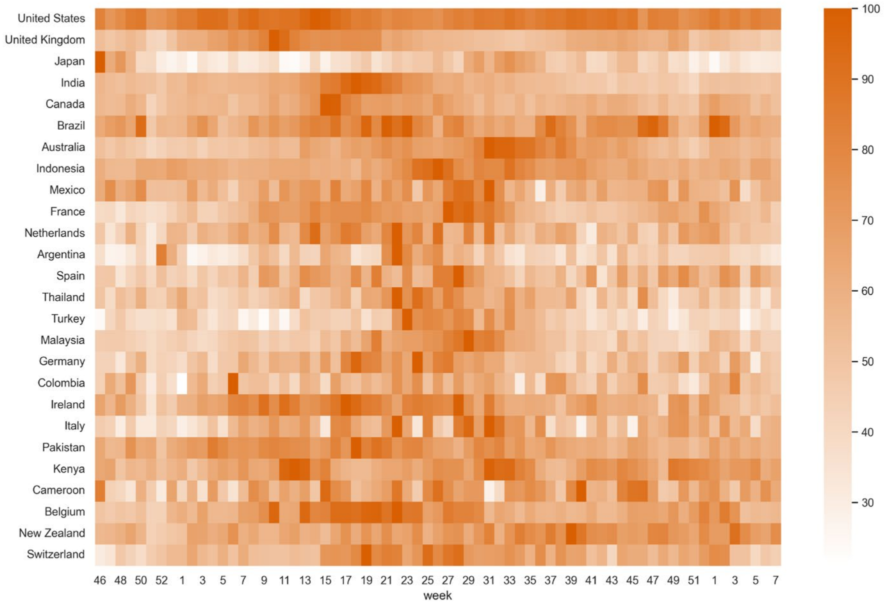

### Coagulation

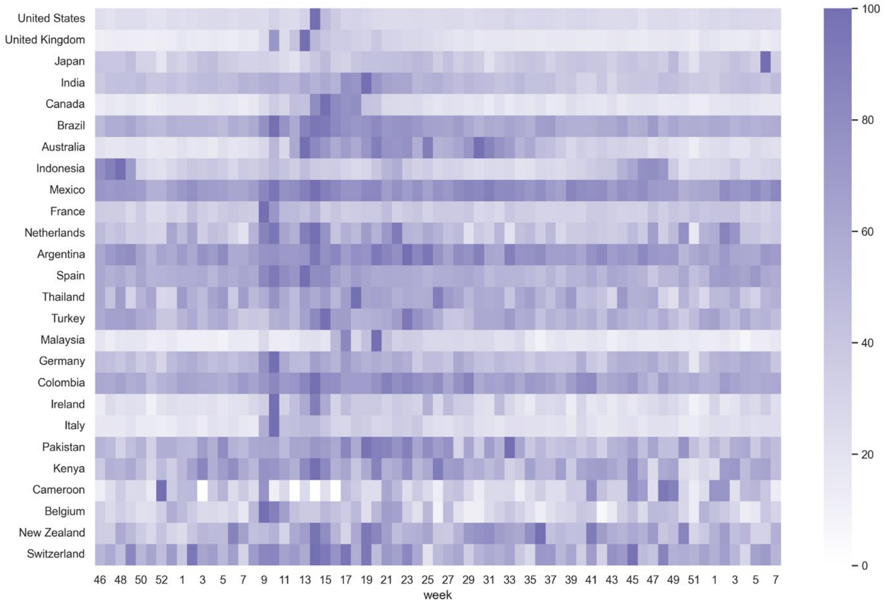

### Fever

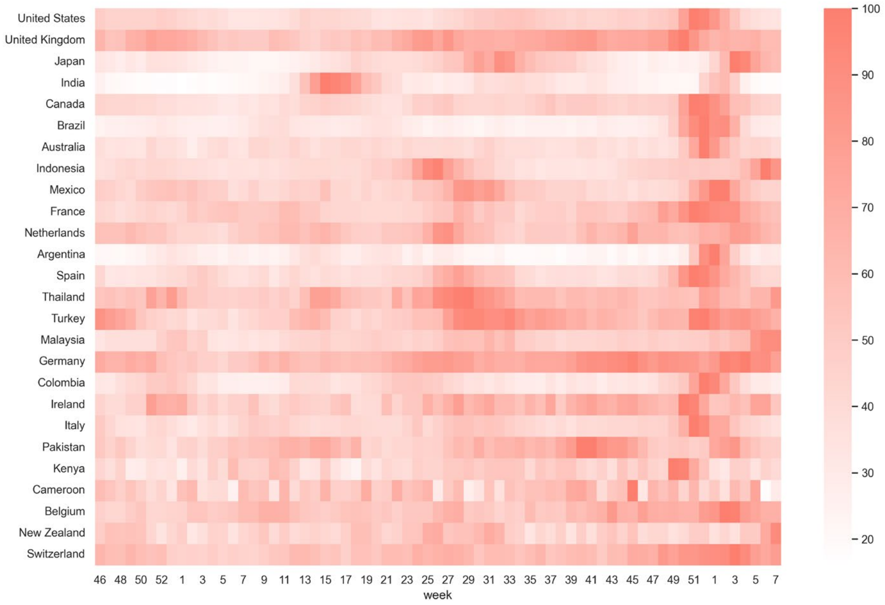

### Guillain–Barré Syndrome

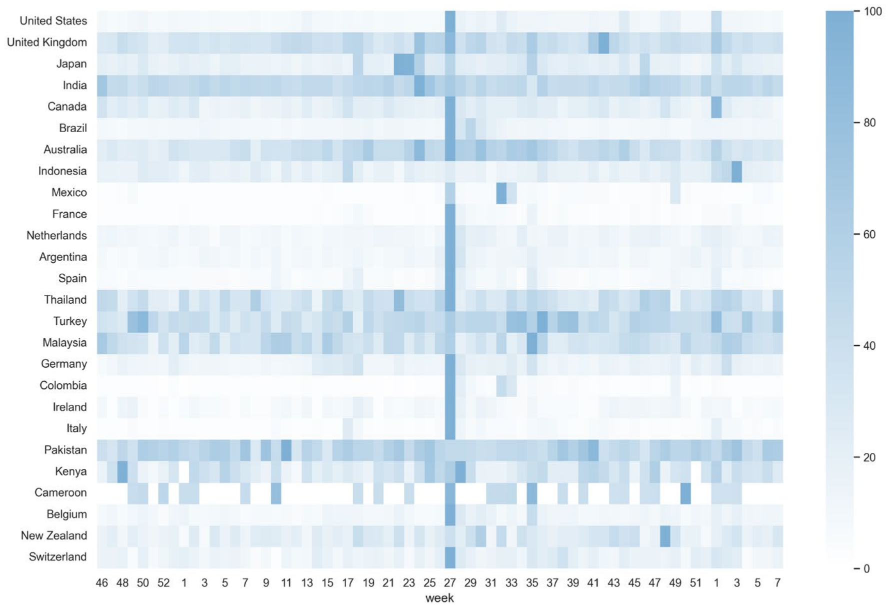

### Myocarditis

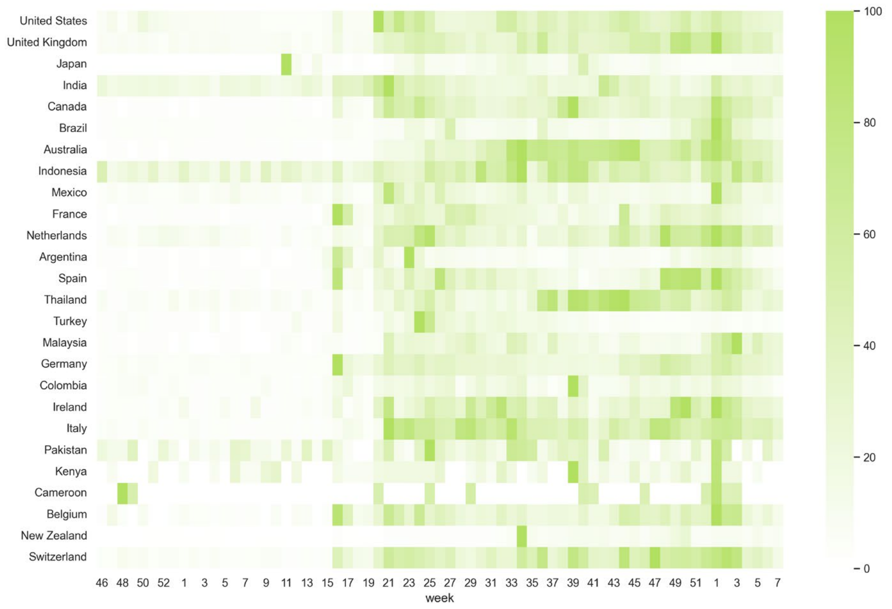

### Pericarditis

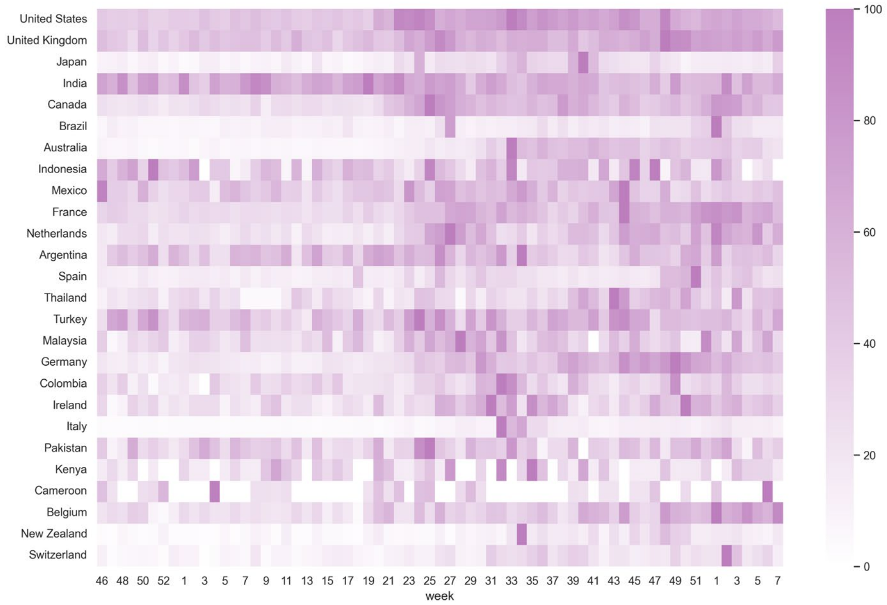

### Side effect

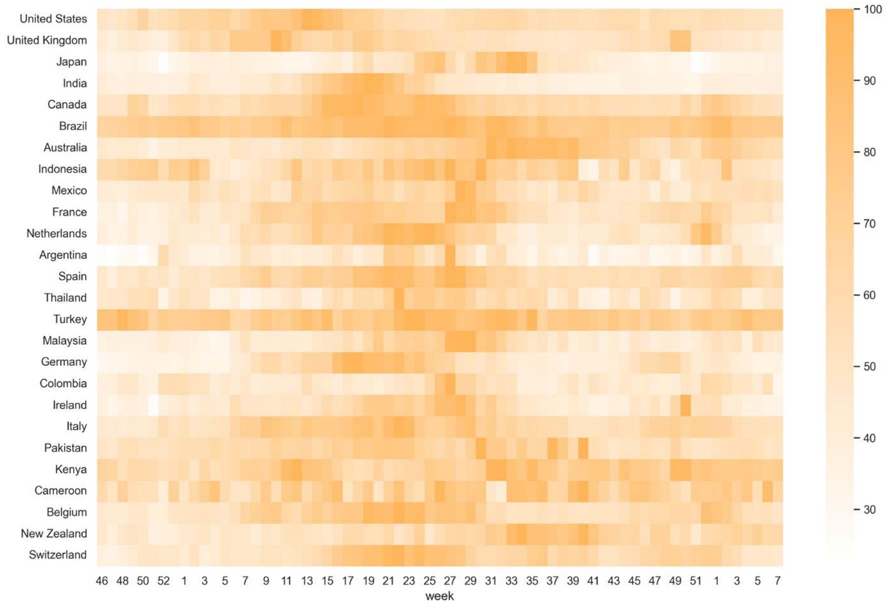

### Skin rash

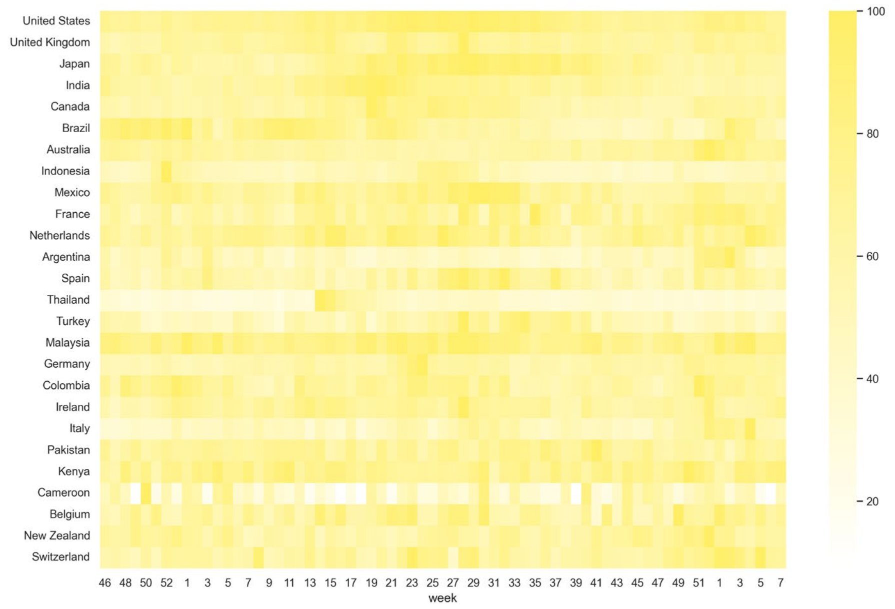

### Thrombosis

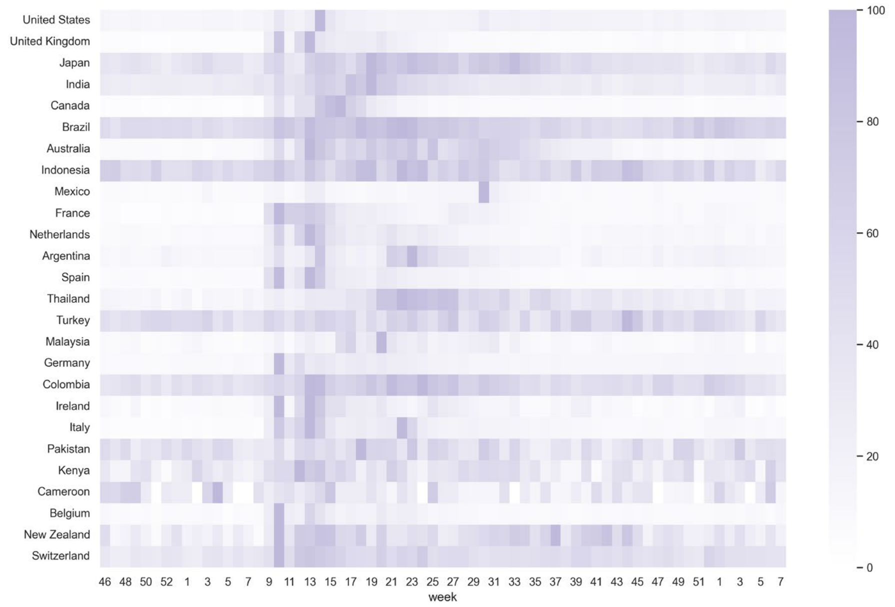

### Thrombus

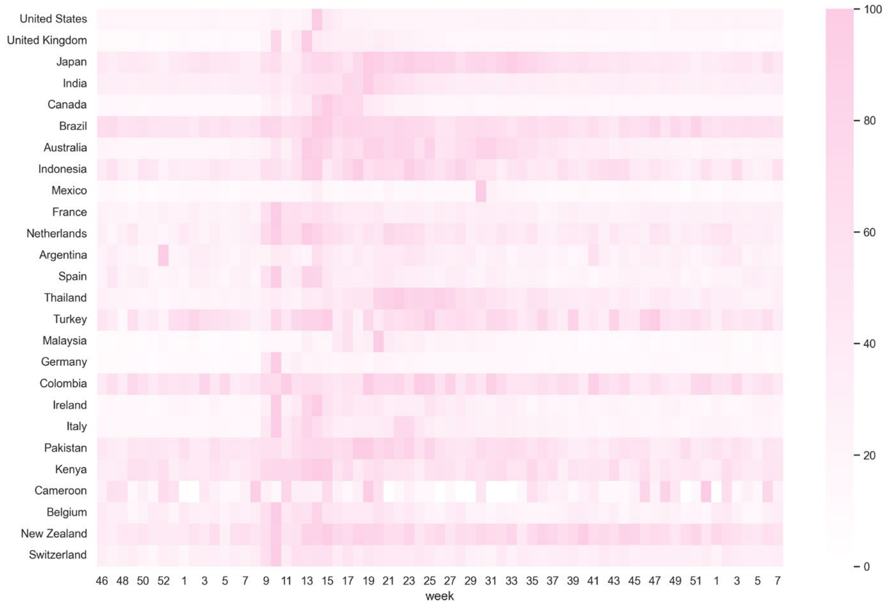

### Vaccine adverse event

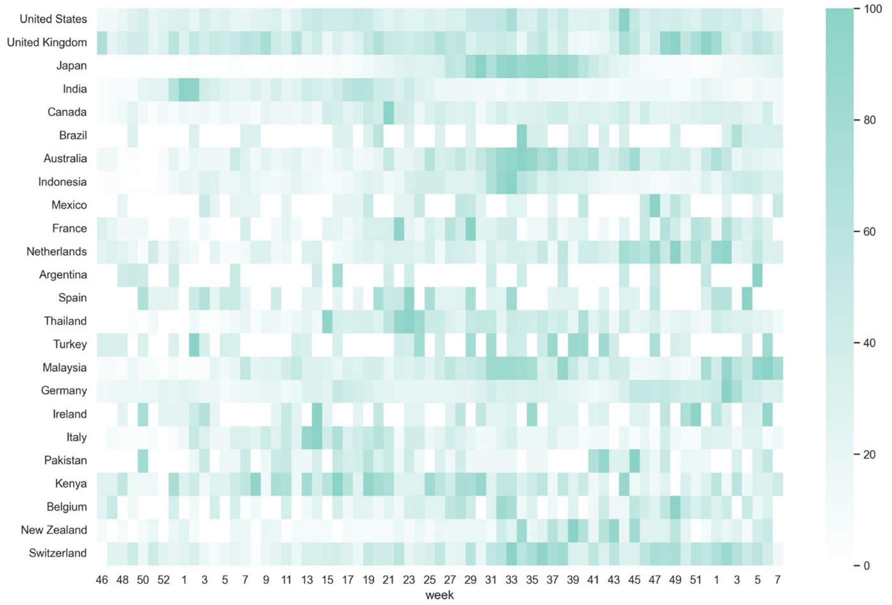
